# Scalable cardiovascular risk assessment using artificial intelligence-enabled event adjudication and widely available hematologic predictors

**DOI:** 10.1101/2021.01.12.21249662

**Authors:** James G. Truslow, Shinichi Goto, Max Homilius, Christopher Mow, John M. Higgins, Calum A. MacRae, Rahul C. Deo

## Abstract

**Introduction:** Researchers routinely evaluate novel biomarkers for incorporation into clinical risk models. Although of potential benefit, such emerging markers, which are often costly or not yet commercially available, are unlikely to enable the scalable risk assessment needed for population health strategies. In contrast, the ideal inputs for population approaches would be those already widely available for most patients. We hypothesized that simple hematologic markers, available in an outpatient complete blood count without differential, would be useful to develop risk models for cardiovascular events.

**Methods:** Using routine laboratory measurements as predictors and neural network-based automated event adjudication of 1,072,348 discharge summaries, we developed and validated models for prediction of heart attack, ischemic stroke, heart failure hospitalization, revascularization, and all-cause mortality.

**Results:** Models with hematology indices alone showed Harrell’s concordance index ranging from 0.60–0.80 on an external validation set. Hematology indices added significantly in terms of discrimination and calibration performance compared to models using only demographic data and diagnostic codes for coronary artery disease, heart failure, and ischemic stroke, with the concordance index of resulting models in the range 0.75–0.85 on an external validation set. Predictive features varied by outcome, and included red blood cell, leukocyte, and platelet indices.

**Conclusion:** We conclude that low-cost ubiquitous inputs, if biologically informative, can provide population-level readouts of risk.

## Introduction

Two approaches guide the development of clinical risk models^1^. One strategy focuses on evaluating innovative markers, typically biomolecular, in a research cohort where banked historical samples and adjudicated outcomes are available. Recent biomarker candidates include genetic variants^2–4^, protein biomarkers^5^, somatic mutations^6^ and serum metabolites^7^. The motivation for such an approach is to improve model performance, while proposing something new about disease mechanisms. One challenge for such a strategy is the lack of availability of such markers in other cohorts, making replication a challenge. The problem of predictor availability extends to model deployment, especially when considering the use of such models to guide population health initiatives. Moreover, given that contributors to disease may evolve over time, insights from a historical cohort may not be readily transportable to new settings^8^.

An alternative approach involves training models using existing low-cost data readily available on a high percentage of patients within a healthcare system. Such an approach is pragmatic and exploits model training as the first step in a systematic program to enable risk assessment for the maximal number of individuals. The output of the models could then be used to guide population health initiatives, hospital- or payer-level financial planning, or clinical trial enrollment^9^. The use of widely available inputs also facilitates iteratively updating models to match changing environments.

In this work, we focus on the latter approach, but use an unconventional choice of predictors: hematologic indices available in a complete blood count without differential (CBC). Moreover, in the spirit of enabling ease of re-training models in new settings, we use a machine-learning strategy for adjudication of acute coronary syndrome, coronary revascularization, heart failure hospitalization, and ischemic stroke, using discharge summaries from hospitalizations^10^. We describe discrimination and calibration performance of the models across two institutions.

## Methods

### Study design and source data

All patients in the model-derivation cohort had a non-urgent outpatient CBC recorded on a Sysmex XE-5000 Automated Hematology System at the Massachusetts General Hospital (MGH) during March 2016 - May 2017. Among 494,654 result-log files stored by the XE-5000s during this period, there were 469,543 unique lab-order identifiers. Where identifiers were associated with more than one file, their timestamps varied by less than 3 hours in 99% of cases. We thus interpreted such multiple readouts as mapping to a single blood sample, and we discarded all but the most recent file. We discarded another 26 files which had internally inconsistent data. Among the 469,517 files we retained for later processing, there were no missing data.

For each patient, we also accessed all diagnostic-code data from the Massachusetts General Brigham (MGB) Electronic Data Warehouse (EDW), including admit diagnoses, encounter diagnoses, medical history, and problem lists, as well as all discharge summaries and progress notes. Encounters, diagnostic codes, and notes could be from any of the institutions within MGB network of hospitals. We excluded all tests that occurred within 7 days prior to or 30 days following an emergency department visit or hospital admission, as defined by the date of a discharge summary note, so that the hematologic indices would more closely reflect the patient’s baseline values.

From the remaining CBCs for each patient, only the earliest one was included, and this CBC defined the patient’s time of entry into the survival study (see below). We excluded patients younger than 30 years at the time of entry into study. Follow-up time was defined from the date of the CBC to the last encounter or note within the MGB-EDW under the following categories: encounters coded as “office visit”, “system generated”, or “hospital encounter”; notes coded as “progress note”, “telephone encounter”, “assessment & plan”, “discharge summary”, “ED provider note”, “consult”, “MR AVS snapshot”, “plan of care”, “patient instruction”, “H&P”. Because of the dramatic change in hospital population caused by COVID-19 in the winter of 2020, we did not consider follow-up or events after 2019 Dec. 31. When a patient’s follow-up extended past this date or if a study outcome occurred after this date, we right censored the patient at 2020 Jan. 1. Patients with no follow-up in the MGB-EDW system were excluded.

The validation cohort consisted of all individuals who had a 10-parameter complete blood count (described below) collected at Brigham Women’s Hospital (BWH) between June 2015 and December 2016, and whose results were available within the MGB-EDW. As with the training data, we excluded laboratory values occurring close in time to emergency department or hospital discharges.

### Artificial intelligence-enabled adjudication of cardiovascular outcomes

We recently trained neural network-based models to classify discharge summaries as associated with one the following four cardiovascular events: acute coronary syndromes (ACS), ischemic stroke (IS), heart-failure hospitalization (HF), and percutaneous coronary intervention/coronary artery bypass surgery (PCI/CABG)^10^. Briefly, after manually labeling 1,372 training notes + 592 validation notes + 1,003 testing notes across 17 institutions, we trained and validated models for event adjudication, with AUROC on the 1,003 test notes as follows: ACS, 0.967; HF, 0.965; IS, 0.980; PCI/CABG, 0.998. The fraction of all discharges in our test set which received labels for CVD events was as follows: ACS, 0.054; HF, 0.122; IS, 0.059; PCI/CABG, 0.054. The four types of events were not mutually exclusive, and a discharge summary could receive as many as four positive labels. For the current work, we selected classification thresholds to maximize *F*_*1*_ score (**Supplementary Table 1A**). The four event models were deployed on the 1,072,348 discharge summaries for the derivation and validation cohorts and a binary event status was determined by each model for each note.

The four AI-adjudicated clinical events, plus all-cause mortality, constituted the five primary outcomes for our survival analysis. In addition to these primary outcomes, we also defined two composite outcomes: 1) acute coronary syndrome or heart-failure hospitalization or ischemic stroke (“ACS/HF/IS”); and 2) ACS/HF/IS or death (“ACS/HF/IS/death”). Date of death was obtained from MGB-EDW, without mediation by statistical methods. We did not employ competing-risk models or exclude a patient from the dataset for outcome *X* if they also experienced outcome *Y*.

Prior history of CAD, IS, or HF was established using diagnostic codes, which we previously evaluated for performance on a set of manually labeled test notes^10^ (**Supplementary Table 1B)**. In contrast to the event models above, which map a single discharge summary to the probability of an event immediately associated with that hospital discharge, ICD codes were taken in aggregate across a patient’s history, to generate a binary status of extant disease up to a certain date.

### Predictors

Predictors used in the sex-specific survival models were:

- Patient’s age at entry into study.
- Binary diagnostic codes for history of each of three conditions, at time of entry into study: heart failure; ischemic stroke; coronary artery disease or myocardial infarction.
- Ten hematologic indices obtained as routine values using the Sysmex XE-5000, at time of entry into study: hematocrit, whole-blood hemoglobin concentration, mean corpuscular hemoglobin, mean corpuscular hemoglobin concentration, mean corpuscular volume, platelet count, red blood cell count, red blood cell distribution width, mean platelet volume, white cell count.

All predictors were centered and scaled except the three binary flags for disease. Some predictor sets included 1^st^ order interactions with age. When main effects were centered and scaled, this transformation was applied before computing interaction terms **(Supplementary Table 3)**.

### Derivation and validation of the models

For each of the seven outcomes, we used *L*_*1*_-penalized Cox proportional hazards models to estimate coefficients for each risk factor. For each outcome, we developed separate models for male and female patients. Penalty strength for each model was selected by optimizing Harrell’s concordance index (C-index)^11^ through *k*-fold cross validation (3 ≤ k ≤ 5). Ties were handled with Breslow’s method. Regression and penalty tuning were performed with the python package scikit-survival v0.14.0^12^.

Each combination of outcome and sex was treated with five different models, each supported by a different set of predictors: set 1 consisted of the ten hematologic indices (“HEM”); set 2 added age to set 1 (“HEM-AGE”); set 3 added disease history to set 2 (“HEM-AGE-HX”); set 4 consisted of age and history (“AGE-HX”); set 5 consisted of just age (“AGE”). Whenever we included age in a predictor set, we also include first-order interactions between age and all main effects **(Supplementary Table 3B)**.

To evaluate performance within the derivation set and provide a measure of uncertainty for the performance estimate, we repeated rounds of 5-fold cross validation, placing the penalty-tuning loop within the model-derivation step for each fold. We then trained final models on the entire derivation dataset. We externally validated each final model on the BWH dataset, having preprocessed the validation data in the same way as the training data. We removed any patients from the validation dataset who also appeared in the derivation dataset.

### Evaluation of model performance

Each combination of outcome, predictors, and sex were evaluated by C-index and Brier score. Brier score was calculated using survival at 3 years post-CBC, using the IPCW method to address censoring^**13**,**14**^. Performance on the derivation dataset is presented with means and confidence intervals computed from repeated *k*-fold cross-validation (40 repeats of 5-fold cross validation). Performance on the validation dataset is presented with means and confidence intervals computed from bootstrapping the validation dataset (1000 bootstrapped samples). Coefficients for final models are estimated using the entire derivation dataset.

Where the performance of two different predictor sets is compared for a single event model (e.g., compare predictor sets HEM-AGE-HX and AGE-HX, for the time-to-death model on the female validation cohort), the comparison is reported as the difference between model scores, as evaluated on a given set of patients. For the derivation dataset, this means that model scores are computed during each fold of cross-validation and compared for the patients in that fold. For the validation dataset, this means that model scores are computed on each bootstrapped sample and compared for the patients in that sample. Thus, a distribution of comparisons is compiled for each type of performance metric.

We evaluate model calibration graphically by plotting deciles of observed event probability vs. predicted probability. Comparisons are made at 3 years of follow-up. Baseline hazard is estimated nonparametrically, using Breslow’s method^15^, and represents the hazard for a patient whose predictors are all zero. This is a patient with average hematologic indices, average age, and with no history of CAD or MI or heart failure or stroke.

### Random Survival Forest

To explore the benefit of modeling high dimensional interactions, we employed an ensemble learning method, random survival forest (RSF)^16^ to train models for each combination of outcome, sex, and predictor set with the help of the Python package scikit-survival v0.14.0. We performed a single round of hyperparameter tuning using *k*-fold cross-validation (k = 5). Hyperparameters optimized included: trees per forest {50,150}; maximum depth of any tree {2,4,6,8}; number of features examined for splitting at each node {1,2,3,4}. We report the mean test-set C-index calculated over the *k* folds. The hyperparameter set represented by the reported C-index is the one that produced the highest mean C-index.

## Results

### Baseline characteristics

The derivation cohort included 11,056 males and 12,182 females, all of whom received at least one outpatient, non-urgent CBC between 2016-Apr-2 and 2017-May-16 (**Table 1A, 1B**). The external validation cohort, at a second institution, consisted of 10771 males and 18900 females, all of whom received at least one outpatient elective CBC between 2015-Jun-1 and 2017-Jan-1. All patients in this study were at least 30 years old at the time the CBC was performed. Unlike other risk-model efforts^17,18^, which have focused on a population free of CVD events and not taking statins, our focus was on a broader set of patients, likely at higher risk. Our rationale was that such individuals are responsible for high resource healthcare utilization, and so could benefit from triggered therapeutic innovations to reduce risk, whether they be novel medications or population-health initiatives.

**Table 1A.** Baseline Demographic Characteristics of Derivation and Validation Cohorts.

| Parameter | Men |  |  | Women |  |  |
| --- | --- | --- | --- | --- | --- | --- |
|  | Deriv. Cohort | Valid. Cohort | *p* | Deriv. Cohort | Valid. Cohort | *p* |
| N | 11056 | 10771 |  | 12182 | 18900 | - |
| Age, years (SD) | 60.1 (14.3) | 59.9 (13.9) | 3.89E-01 | 57.4 (15.0) | 53.7 (15.4) | 4.17E-95 |
| Race |  |  |  |  |  |  |
| White, % (N) | 79.2 (8756) | 80.3 (8652) | 9.27E-67 | 78.0 (9497) | 73.1 (13808) | 1.52E-107 |
| Black or African American, % (N) | 5.5 (606) | 6.6 (714) |  | 5.7 (698) | 9.9 (1872) |  |
| Hispanic or Latino, % (N) | 0.4 (45) | 2.8 (303) |  | 0.9 (107) | 4.1 (779) |  |
| Asian, % (N) | 4.9 (543) | 3.3 (356) |  | 5.5 (664) | 5.5 (1042) |  |
| American Indian or Alaska Native, % (N) | 0.1 (8) | 0.2 (19) |  | 0.1 (12) | 0.1 (18) |  |
| Native Hawaiian or Other Pacific Islander, % (N) | 0.0 (3) | 0.1 (10) |  | 0.1 (7) | 0.0 (6) |  |
| Missing, % (N) | 9.9 (1095) | 6.7 (717) |  | 9.8 (1197) | 7.3 (1375) |  |
| Hispanic Origin |  |  |  |  |  |  |
| Hispanic, % (N) | 4.8 (536) | 5.8 (628) | 2.89E-64 | 6.5 (789) | 8.5 (1614) | 2.57E-74 |
| Non-Hispanic, % (N) | 85.0 (9399) | 76.3 (8222) |  | 84.2 (10252) | 75.7 (14301) |  |
| Missing, % (N) | 10.1 (1121) | 17.8 (1921) |  | 9.4 (1141) | 15.8 (2985) |  |
| Smoking |  |  |  |  |  |  |
| Current, % (N) | 8.0 (887) | 6.9 (743) | 1.07E-50 | 5.7 (690) | 4.7 (881) | 5.34E-41 |
| Former, % (N) | 30.7 (3393) | 34.9 (3757) |  | 25.7 (3133) | 26.5 (5000) |  |
| Never, % (N) | 49.3 (5454) | 51.9 (5591) |  | 58.0 (7071) | 62.4 (11789) |  |
| Missing, % (N) | 12.0 (1322) | 6.3 (680) |  | 10.6 (1288) | 6.5 (1230) |  |

Significance for age difference is via *t*-test. The other three differences are compared via $\chi^2$ tests.

**Table 1B.** Baseline Clinical Characteristics of Derivation and Validation Cohorts.

| Parameter | Men |  |  | Women |  |  |
| --- | --- | --- | --- | --- | --- | --- |
|  | Deriv. Cohort | Valid. Cohort | *p* | Deriv. Cohort | Valid. Cohort | *p* |
| Heart rate, bpm, mean (SD, N<sub>MISS</sub>) | 73.6 (11.5, 1171) | 72.5 (11.8, 222) | 2.7E-12 | 76.5 (11.0, 1375) | 75.8 (11.1, 812) | 2.5E-08 |
| Systolic BP, mmHg, mean (SD, N<sub>MISS</sub>) | 128.0 (13.4, 1092) | 129.2 (13.5, 214) | 3.7E-10 | 123.9 (14.3, 1170) | 123.8 (14.7, 246) | 5.1E-01 |
| Diastolic BP, mmHg, mean (SD, N<sub>MISS</sub>) | 75.3 (8.4, 1092) | 75.3 (9.7, 214) | 8.2E-01 | 73.3 (7.9, 1170) | 70.9 (8.5, 246) | 4E-127 |
| BMI kg/m<sup>2</sup>, median (IQR, N<sub>MISS</sub>) | 27.9 (25.1-31.5, 1759) | 28.0 (25.3-31.5, 530) | 8.5E-01 | 26.4 (22.7-31.3, 1811) | 26.5 (22.9-31.4, 1200) | 1.3E-01 |
| LDL mg/dL, mean (SD, N<sub>MISS</sub>) | 96.6 (35.5, 8221) | 99.2 (33.3, 3128) | 8.3E-04 | 109.1 (35.9, 9529) | 106.4 (32.6, 7359) | 2.6E-04 |
| HDL mg/dL, mean (SD, N<sub>MISS</sub>) | 47.8 (15.2, 4665) | 51.6 (16.3, 3037) | 3.6E-46 | 61.0 (19.4, 5918) | 66.5 (20.3, 7143) | 2.6E-70 |
| Triglycerides, mg/dL, mean (SD, N<sub>MISS</sub>) | 131.3 (96.6, 4660) | 137.9 (98.5, 3003) | 5.2E-05 | 115.6 (72.8, 5950) | 115.9 (73.4, 7155) | 8.0E-01 |
| eGFR mL/min/1.73m<sup>2</sup>, mean (SD, N<sub>MISS</sub>) | 67.1 (23.2, 2322) | 67.3 (21.1, 417) | 4.3E-1 | 91.0 (26.7, 3140) | 94.7 (24.8, 2564) | 6.4E-28 |
| Ejection fraction, %, mean (SD, N<sub>MISS</sub>) | 59.6 (14.8, 10130) | 56.6 (12.3, 9192) | 2.7E-07 | 64.3 (11.1, 11448) | 61.1 (8.6, 17319) | 1.4E-11 |
| Coronary artery disease, % (N) | 18.3 (2026) | 16.1 (1739) | 2.0E-05 | 6.8 (824) | 4.1 (777) | 5.1E-25 |
| Heart failure, % (N) | 9.3 (1028) | 8.4 (909) | 2.6E-02 | 5.0 (613) | 3.3 (619) | 9.2E-15 |
| Ischemic stroke, % (N) | 3.6 (403) | 3.2 (349) | 1.0E-01 | 2.5 (309) | 1.7 (328) | 1.1E-06 |
| Type 2 diabetes mellitus, % (N) | 18.8 (2083) | 17.7 (1907) | 3.0E-02 | 11.5 (1398) | 10.8 (2032) | 4.6E-02 |
| Hypertension, % (N) | 51.4 (5678) | 51.0 (5488) | 5.5E-01 | 38.5 (4691) | 34.2 (6460) | 8.1E-15 |
| Atrial fibrillation, % (N) | 10.9 (1210) | 11.8 (1272) | 4.4E-02 | 5.1 (617) | 3.9 (734) | 6.2E-07 |
| Aspirin, % (N) | 44.2 (4888) | 43.4 (4673) | 2.2E-01 | 27.0 (3289) | 22.9 (4329) | 2.6E-16 |
| Statin, % (N) | 45.7 (5057) | 46.4 (5002) | 3.0E-01 | 27.7 (3369) | 23.8 (4493) | 1.5E-14 |
| ACE inhibitor, % (N) | 23.7 (2615) | 26.0 (2798) | 7.0E-05 | 13.7 (1664) | 12.7 (2400) | 1.4E-02 |
| Angiotensin receptor blocker, % (N) | 12.7 (1401) | 12.5 (1342) | 6.4E-01 | 9.9 (1204) | 8.8 (1665) | 1.4E-03 |
| Beta blocker, % (N) | 32.9 (3634) | 34.2 (3685) | 3.6E-02 | 23.4 (2853) | 19.5 (3684) | 1.1E-16 |

Means are compared via *t*-test. Proportions are compared via Z-test.

Median follow-up time for the derivation cohort was 1,091 days, and for the validation cohort was 1,517 days (**Supplementary Figures 1 and 2**). The most notable differences between derivation and validation cohorts were a younger age in the female validation cohort compared to the derivation cohort (53.7 vs. 57.4 years) and greater racial diversity within the validation cohorts (e.g., 19.6% non-White in female validation vs. 12.3% in female derivation). Prevalence of cardiovascular disease in the male derivation cohort was similar to that in the male validation cohort, whereas prevalence in the female derivation cohort was higher than in the validation cohort.

CONSORT-type diagrams showing the flow of patients and their data through our analysis are available in the Supplement (**Supplementary Figures 3 and 4)**.

### Rates of cardiovascular disease and death

In the derivation cohort, the proportion of patients with one or more ACS events within the follow-up period was 0.019, with one or more HF hospitalizations was 0.046, with one or more ischemic strokes was 0.022, with one or more coronary revascularizations was 0.023 (**Tables 2A, 2B**). The fraction who died was 0.054. In the validation cohort, the proportion of patients with one or more ACS events was 0.013, with one or more HF hospitalizations was 0.045, with one or more ischemic strokes was 0.021, with one or more coronary revascularizations was 0.014. The fraction who died was 0.050.

**Table 2A.** Case Count in Derivation Cohort.

| Outcome | Age | Women |  |  | Men |  |  |
| --- | --- | --- | --- | --- | --- | --- | --- |
|  |  | Cases | Total | Cases/1000 | Cases | Total | Cases/1000 |
| ACS | <45 | 3 | 2917 | 1.0 (-0.1 to 2.2) | 11 | 1814 | 6.1 (2.5 to 9.6) |
|  | 45-60 | 21 | 3828 | 5.5 (3.1 to 7.8) | 73 | 3475 | 21.0 (16.2 to 25.8) |
|  | >60 | 88 | 5432 | 16.2 (12.8 to 19.6) | 247 | 5759 | 42.9 (37.7 to 48.1) |
| HF | <45 | 39 | 2917 | 13.4 (9.2 to 17.5) | 31 | 1814 | 17.1 (11.1 to 23.1) |
|  | 45-60 | 75 | 3828 | 19.6 (15.2 to 24.0) | 146 | 3475 | 42.0 (35.3 to 48.7) |
|  | >60 | 327 | 5432 | 60.2 (53.9 to 66.5) | 459 | 5759 | 79.7 (72.7 to 86.7) |
| IS | <45 | 14 | 2917 | 4.8 (2.3 to 7.3) | 16 | 1814 | 8.8 (4.5 to 13.1) |
|  | 45-60 | 32 | 3828 | 8.4 (5.5 to 11.2) | 60 | 3475 | 17.3 (12.9 to 21.6) |
|  | >60 | 187 | 5432 | 34.4 (29.6 to 39.3) | 206 | 5759 | 35.8 (31.0 to 40.6) |
| PCI/<br>CABG | <45 | 6 | 2917 | 2.1 (0.4 to 3.7) | 8 | 1814 | 4.4 (1.4 to 7.5) |
|  | 45-60 | 23 | 3828 | 6.0 (3.6 to 8.5) | 88 | 3475 | 25.3 (20.1 to 30.5) |
|  | >60 | 106 | 5432 | 19.5 (15.8 to 23.2) | 294 | 5759 | 51.1 (45.4 to 56.7) |
| Death | <45 | 22 | 2917 | 7.5 (4.4 to 10.7) | 21 | 1815 | 11.6 (6.7 to 16.5) |
|  | 45-60 | 94 | 3831 | 24.5 (19.6 to 29.4) | 122 | 3476 | 35.1 (29.0 to 41.2) |
|  | >60 | 414 | 5434 | 76.2 (69.1 to 83.2) | 575 | 5765 | 99.7 (92.0 to 107.5) |

**Table 2B.**
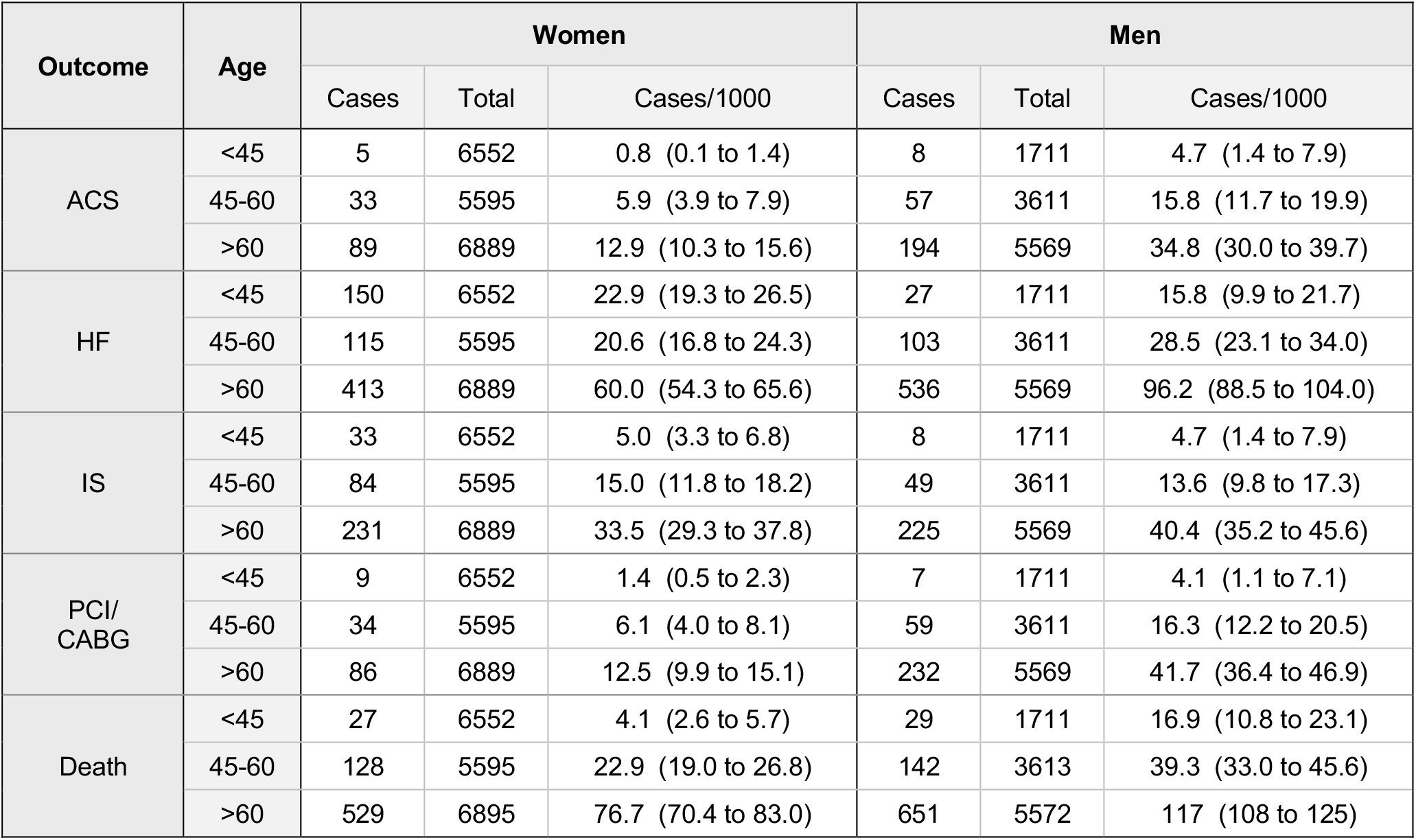
Case Count in Validation Cohort. Number of patients in **(A)** derivation and **(B)** validation cohorts, and number of each type of outcome observed, as well as the fraction of patients with each outcome. Fraction is reported as outcomes per 1000 patients, along with 95% confidence intervals for the fraction.

| Outcome | Age | Women |  |  | Men |  |  |
| --- | --- | --- | --- | --- | --- | --- | --- |
|  |  | Cases | Total | Cases/1000 | Cases | Total | Cases/1000 |
| ACS | <45 | 5 | 6552 | 0.8 (0.1 to 1.4) | 8 | 1711 | 4.7 (1.4 to 7.9) |
|  | 45-60 | 33 | 5595 | 5.9 (3.9 to 7.9) | 57 | 3611 | 15.8 (11.7 to 19.9) |
|  | >60 | 89 | 6889 | 12.9 (10.3 to 15.6) | 194 | 5569 | 34.8 (30.0 to 39.7) |
| HF | <45 | 150 | 6552 | 22.9 (19.3 to 26.5) | 27 | 1711 | 15.8 (9.9 to 21.7) |
|  | 45-60 | 115 | 5595 | 20.6 (16.8 to 24.3) | 103 | 3611 | 28.5 (23.1 to 34.0) |
|  | >60 | 413 | 6889 | 60.0 (54.3 to 65.6) | 536 | 5569 | 96.2 (88.5 to 104.0) |
| IS | <45 | 33 | 6552 | 5.0 (3.3 to 6.8) | 8 | 1711 | 4.7 (1.4 to 7.9) |
|  | 45-60 | 84 | 5595 | 15.0 (11.8 to 18.2) | 49 | 3611 | 13.6 (9.8 to 17.3) |
|  | >60 | 231 | 6889 | 33.5 (29.3 to 37.8) | 225 | 5569 | 40.4 (35.2 to 45.6) |
| PCI/<br>CABG | <45 | 9 | 6552 | 1.4 (0.5 to 2.3) | 7 | 1711 | 4.1 (1.1 to 7.1) |
|  | 45-60 | 34 | 5595 | 6.1 (4.0 to 8.1) | 59 | 3611 | 16.3 (12.2 to 20.5) |
|  | >60 | 86 | 6889 | 12.5 (9.9 to 15.1) | 232 | 5569 | 41.7 (36.4 to 46.9) |
| Death | <45 | 27 | 6552 | 4.1 (2.6 to 5.7) | 29 | 1711 | 16.9 (10.8 to 23.1) |
|  | 45-60 | 128 | 5595 | 22.9 (19.0 to 26.8) | 142 | 3613 | 39.3 (33.0 to 45.6) |
|  | >60 | 529 | 6895 | 76.7 (70.4 to 83.0) | 651 | 5572 | 117 (108 to 125) |

### Model performance

Model performance was assessed in the derivation cohort using repeated 5-fold cross validation (**Table 3**). We refer to the held-out data defined by repeated *k*-fold cross validation as the internal test set (though, to be clear, the process of repeated *k*-fold cross validation operated on 100% of the derivation cohort). Results from the external validation cohort are obtained by the bootstrap method ^19^.

**Table 3.**
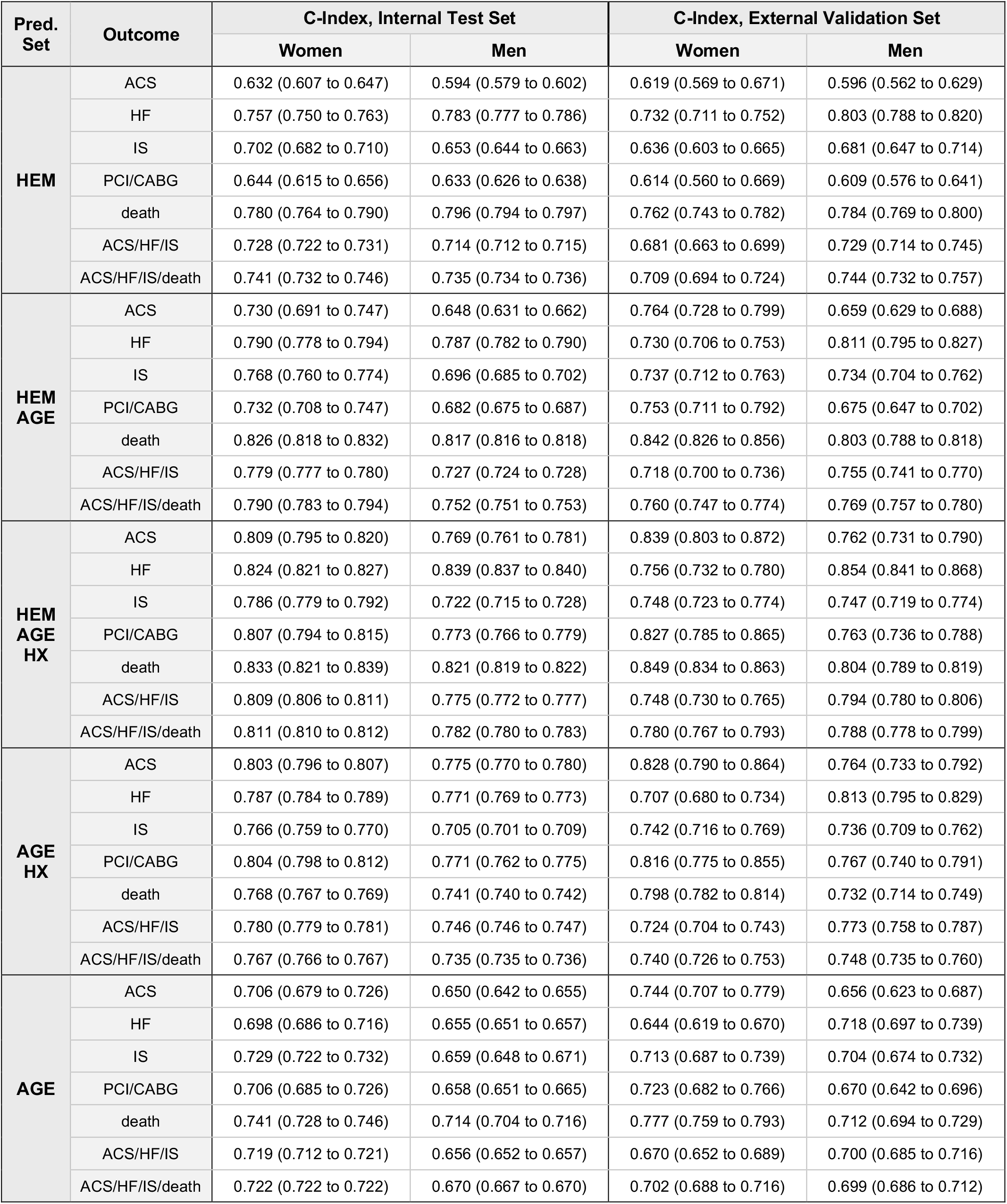
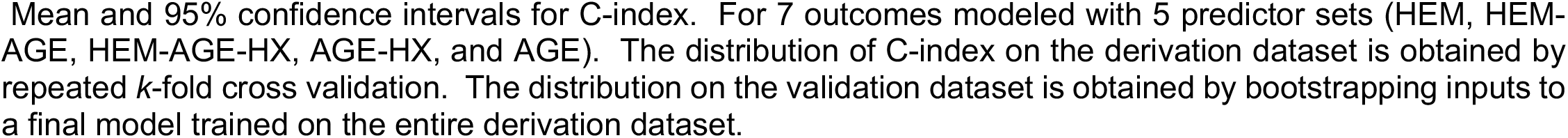
Model performance, according to C-index.

| Pred. Set | Outcome | C-Index, Internal Test Set |  | C-Index, External Validation Set |  |
| --- | --- | --- | --- | --- | --- |
|  |  | Women | Men | Women | Men |
| HEM | ACS | 0.632 (0.607 to 0.647) | 0.594 (0.579 to 0.602) | 0.619 (0.569 to 0.671) | 0.596 (0.562 to 0.629) |
|  | HF | 0.757 (0.750 to 0.763) | 0.783 (0.777 to 0.786) | 0.732 (0.711 to 0.752) | 0.803 (0.788 to 0.820) |
|  | IS | 0.702 (0.682 to 0.710) | 0.653 (0.644 to 0.663) | 0.636 (0.603 to 0.665) | 0.681 (0.647 to 0.714) |
|  | PCI/CABG | 0.644 (0.615 to 0.656) | 0.633 (0.626 to 0.638) | 0.614 (0.560 to 0.669) | 0.609 (0.576 to 0.641) |
|  | death | 0.780 (0.764 to 0.790) | 0.796 (0.794 to 0.797) | 0.762 (0.743 to 0.782) | 0.784 (0.769 to 0.800) |
|  | ACS/HF/IS | 0.728 (0.722 to 0.731) | 0.714 (0.712 to 0.715) | 0.681 (0.663 to 0.699) | 0.729 (0.714 to 0.745) |
|  | ACS/HF/IS/death | 0.741 (0.732 to 0.746) | 0.735 (0.734 to 0.736) | 0.709 (0.694 to 0.724) | 0.744 (0.732 to 0.757) |
| HEM AGE | ACS | 0.730 (0.691 to 0.747) | 0.648 (0.631 to 0.662) | 0.764 (0.728 to 0.799) | 0.659 (0.629 to 0.688) |
|  | HF | 0.790 (0.778 to 0.794) | 0.787 (0.782 to 0.790) | 0.730 (0.706 to 0.753) | 0.811 (0.795 to 0.827) |
|  | IS | 0.768 (0.760 to 0.774) | 0.696 (0.685 to 0.702) | 0.737 (0.712 to 0.763) | 0.734 (0.704 to 0.762) |
|  | PCI/CABG | 0.732 (0.708 to 0.747) | 0.682 (0.675 to 0.687) | 0.753 (0.711 to 0.792) | 0.675 (0.647 to 0.702) |
|  | death | 0.826 (0.818 to 0.832) | 0.817 (0.816 to 0.818) | 0.842 (0.826 to 0.856) | 0.803 (0.788 to 0.818) |
|  | ACS/HF/IS | 0.779 (0.777 to 0.780) | 0.727 (0.724 to 0.728) | 0.718 (0.700 to 0.736) | 0.755 (0.741 to 0.770) |
|  | ACS/HF/IS/death | 0.790 (0.783 to 0.794) | 0.752 (0.751 to 0.753) | 0.760 (0.747 to 0.774) | 0.769 (0.757 to 0.780) |
| HEM AGE HX | ACS | 0.809 (0.795 to 0.820) | 0.769 (0.761 to 0.781) | 0.839 (0.803 to 0.872) | 0.762 (0.731 to 0.790) |
|  | HF | 0.824 (0.821 to 0.827) | 0.839 (0.837 to 0.840) | 0.756 (0.732 to 0.780) | 0.854 (0.841 to 0.868) |
|  | IS | 0.786 (0.779 to 0.792) | 0.722 (0.715 to 0.728) | 0.748 (0.723 to 0.774) | 0.747 (0.719 to 0.774) |
|  | PCI/CABG | 0.807 (0.794 to 0.815) | 0.773 (0.766 to 0.779) | 0.827 (0.785 to 0.865) | 0.763 (0.736 to 0.788) |
|  | death | 0.833 (0.821 to 0.839) | 0.821 (0.819 to 0.822) | 0.849 (0.834 to 0.863) | 0.804 (0.789 to 0.819) |
|  | ACS/HF/IS | 0.809 (0.806 to 0.811) | 0.775 (0.772 to 0.777) | 0.748 (0.730 to 0.765) | 0.794 (0.780 to 0.806) |
|  | ACS/HF/IS/death | 0.811 (0.810 to 0.812) | 0.782 (0.780 to 0.783) | 0.780 (0.767 to 0.793) | 0.788 (0.778 to 0.799) |
| AGE HX | ACS | 0.803 (0.796 to 0.807) | 0.775 (0.770 to 0.780) | 0.828 (0.790 to 0.864) | 0.764 (0.733 to 0.792) |
|  | HF | 0.787 (0.784 to 0.789) | 0.771 (0.769 to 0.773) | 0.707 (0.680 to 0.734) | 0.813 (0.795 to 0.829) |
|  | IS | 0.766 (0.759 to 0.770) | 0.705 (0.701 to 0.709) | 0.742 (0.716 to 0.769) | 0.736 (0.709 to 0.762) |
|  | PCI/CABG | 0.804 (0.798 to 0.812) | 0.771 (0.762 to 0.775) | 0.816 (0.775 to 0.855) | 0.767 (0.740 to 0.791) |
|  | death | 0.768 (0.767 to 0.769) | 0.741 (0.740 to 0.742) | 0.798 (0.782 to 0.814) | 0.732 (0.714 to 0.749) |
|  | ACS/HF/IS | 0.780 (0.779 to 0.781) | 0.746 (0.746 to 0.747) | 0.724 (0.704 to 0.743) | 0.773 (0.758 to 0.787) |
|  | ACS/HF/IS/death | 0.767 (0.766 to 0.767) | 0.735 (0.735 to 0.736) | 0.740 (0.726 to 0.753) | 0.748 (0.735 to 0.760) |
| AGE | ACS | 0.706 (0.679 to 0.726) | 0.650 (0.642 to 0.655) | 0.744 (0.707 to 0.779) | 0.656 (0.623 to 0.687) |
|  | HF | 0.698 (0.686 to 0.716) | 0.655 (0.651 to 0.657) | 0.644 (0.619 to 0.670) | 0.718 (0.697 to 0.739) |
|  | IS | 0.729 (0.722 to 0.732) | 0.659 (0.648 to 0.671) | 0.713 (0.687 to 0.739) | 0.704 (0.674 to 0.732) |
|  | PCI/CABG | 0.706 (0.685 to 0.726) | 0.658 (0.651 to 0.665) | 0.723 (0.682 to 0.766) | 0.670 (0.642 to 0.696) |
|  | death | 0.741 (0.728 to 0.746) | 0.714 (0.704 to 0.716) | 0.777 (0.759 to 0.793) | 0.712 (0.694 to 0.729) |
|  | ACS/HF/IS | 0.719 (0.712 to 0.721) | 0.656 (0.652 to 0.657) | 0.670 (0.652 to 0.689) | 0.700 (0.685 to 0.716) |
|  | ACS/HF/IS/death | 0.722 (0.722 to 0.722) | 0.670 (0.667 to 0.670) | 0.702 (0.688 to 0.716) | 0.699 (0.686 to 0.712) |

Mean and 95% confidence intervals for C-index. For 7 outcomes modeled with 5 predictor sets (HEM, HEM-AGE, HEM-AGE-HX, AGE-HX, and AGE). The distribution of C-index on the derivation dataset is obtained by repeated $k$ -fold cross validation. The distribution on the validation dataset is obtained by bootstrapping inputs to a final model trained on the entire derivation dataset.

#### The relationship of age to cardiovascular outcomes: AGE predictor set

Age is a powerful predictor for cardiovascular outcomes. On the internal test set, the AGE models produced C-indices between 0.65 and 0.74, whereas in the external validation cohort, the AGE models produced C-indices between 0.64 and 0.78 (**Table 3**). Brier scores were < 0.15 for all models in this study, their low magnitude due primarily to the low frequency of events in this dataset (**Supplementary Table 4**).

#### The utility of hematologic predictors for predicting cardiovascular events: HEM, and HEM-AGE vs AGE

Hematologic predictors also had utility for predicting cardiovascular outcomes. On the internal test set, C-indices for the HEM models ranged from 0.59 to 0.80 whereas on the validation set, C-indices for the HEM models ranged from 0.61 to 0.80 (**Table 3**). For both cohorts, the best discrimination was seen for heart failure hospitalization, composite cardiovascular outcomes, and death for both sexes, whereas the worst prediction was seen for ACS risk in men.

In the internal test set, when we added hematologic predictors to the AGE models (thus making the HEM-AGE models), we observed superior discrimination for almost all of our outcomes compared to models predicting on age alone (**Table 4**). The only models that did not clearly benefit from adding the hematologic predictors were models for ACS and PCI/CABG, in which cases the improvement in C-index was small or not statistically significant. Out of 14 comparisons (7 outcomes × 2 sexes) for these two predictor sets, 11 showed a statistically significant change in C-index on the internal test set. Of those 11, 10 also showed a corroborating statistically significant change on the external validation set.

**Table 4.**
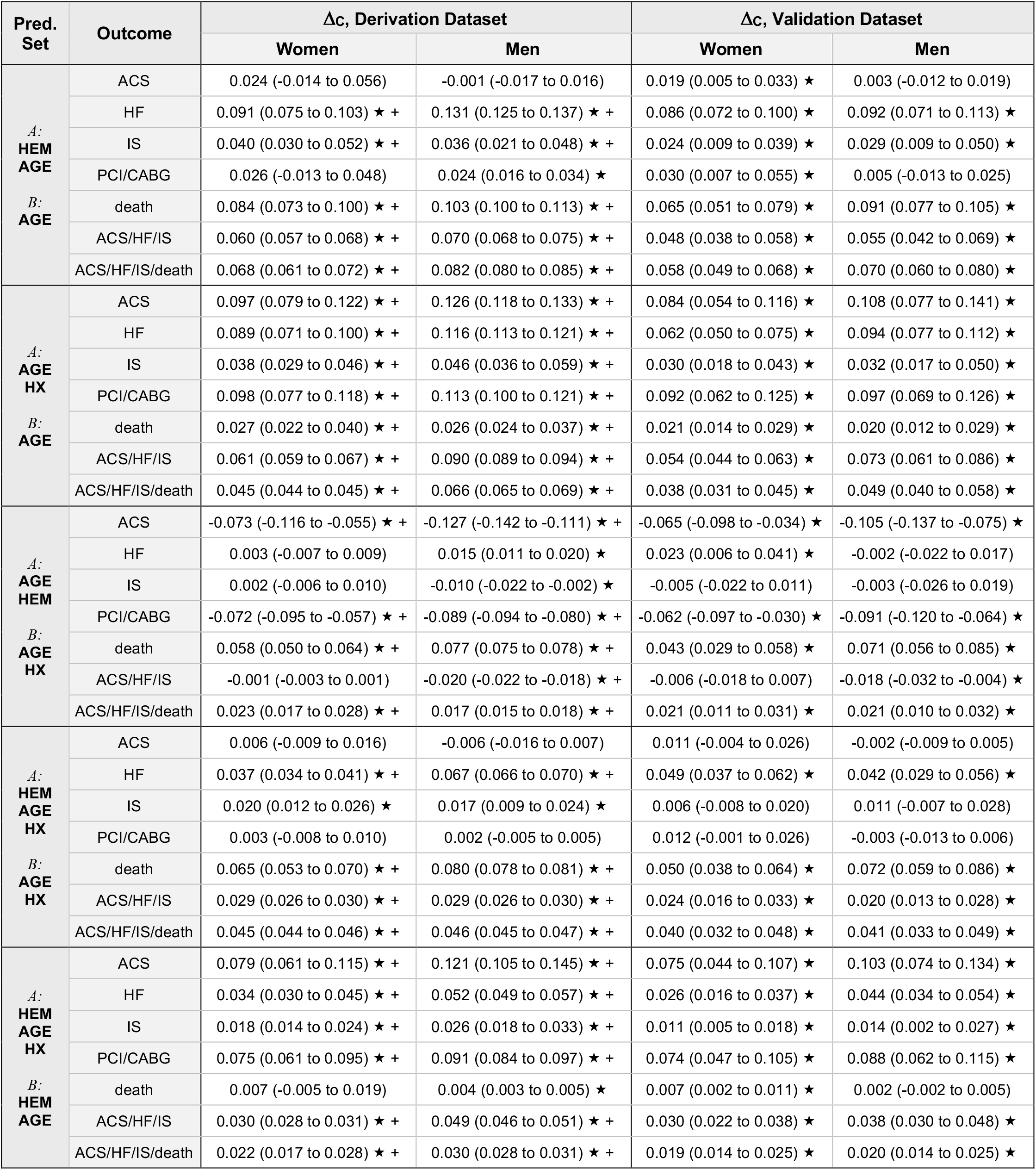
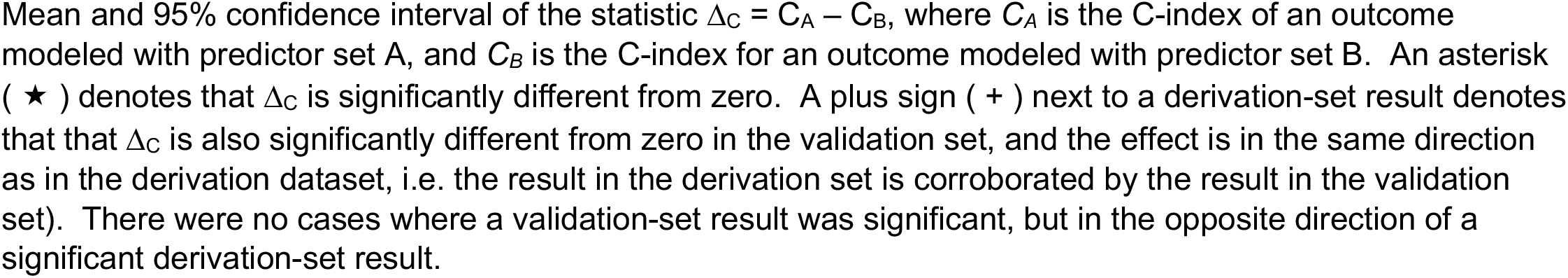
Comparison of pairs of predictor sets according to C-index.

| Pred. Set | Outcome | $\Delta c$, Derivation Dataset | | $\Delta c$, Validation Dataset | |
| --- | --- | --- | --- | --- | --- |
|  |  | Women | Men | Women | Men |
| **A:**<br>HEM<br>AGE<br>**B:**<br>AGE | ACS | 0.024 (-0.014 to 0.056) | -0.001 (-0.017 to 0.016) | 0.019 (0.005 to 0.033) ★ | 0.003 (-0.012 to 0.019) |
|  | HF | 0.091 (0.075 to 0.103) ★ + | 0.131 (0.125 to 0.137) ★ + | 0.086 (0.072 to 0.100) ★ | 0.092 (0.071 to 0.113) ★ |
|  | IS | 0.040 (0.030 to 0.052) ★ + | 0.036 (0.021 to 0.048) ★ + | 0.024 (0.009 to 0.039) ★ | 0.029 (0.009 to 0.050) ★ |
|  | PCI/CABG | 0.026 (-0.013 to 0.048) | 0.024 (0.016 to 0.034) ★ | 0.030 (0.007 to 0.055) ★ | 0.005 (-0.013 to 0.025) |
|  | death | 0.084 (0.073 to 0.100) ★ + | 0.103 (0.100 to 0.113) ★ + | 0.065 (0.051 to 0.079) ★ | 0.091 (0.077 to 0.105) ★ |
|  | ACS/HF/IS | 0.060 (0.057 to 0.068) ★ + | 0.070 (0.068 to 0.075) ★ + | 0.048 (0.038 to 0.058) ★ | 0.055 (0.042 to 0.069) ★ |
|  | ACS/HF/IS/death | 0.068 (0.061 to 0.072) ★ + | 0.082 (0.080 to 0.085) ★ + | 0.058 (0.049 to 0.068) ★ | 0.070 (0.060 to 0.080) ★ |
| **A:**<br>AGE<br>HX<br>**B:**<br>AGE | ACS | 0.097 (0.079 to 0.122) ★ + | 0.126 (0.118 to 0.133) ★ + | 0.084 (0.054 to 0.116) ★ | 0.108 (0.077 to 0.141) ★ |
|  | HF | 0.089 (0.071 to 0.100) ★ + | 0.116 (0.113 to 0.121) ★ + | 0.062 (0.050 to 0.075) ★ | 0.094 (0.077 to 0.112) ★ |
|  | IS | 0.038 (0.029 to 0.046) ★ + | 0.046 (0.036 to 0.059) ★ + | 0.030 (0.018 to 0.043) ★ | 0.032 (0.017 to 0.050) ★ |
|  | PCI/CABG | 0.098 (0.077 to 0.118) ★ + | 0.113 (0.100 to 0.121) ★ + | 0.092 (0.062 to 0.125) ★ | 0.097 (0.069 to 0.126) ★ |
|  | death | 0.027 (0.022 to 0.040) ★ + | 0.026 (0.024 to 0.037) ★ + | 0.021 (0.014 to 0.029) ★ | 0.020 (0.012 to 0.029) ★ |
|  | ACS/HF/IS | 0.061 (0.059 to 0.067) ★ + | 0.090 (0.089 to 0.094) ★ + | 0.054 (0.044 to 0.063) ★ | 0.073 (0.061 to 0.086) ★ |
|  | ACS/HF/IS/death | 0.045 (0.044 to 0.045) ★ + | 0.066 (0.065 to 0.069) ★ + | 0.038 (0.031 to 0.045) ★ | 0.049 (0.040 to 0.058) ★ |
| **A:**<br>AGE<br>HEM<br>**B:**<br>AGE<br>HX | ACS | -0.073 (-0.116 to -0.055) ★ + | -0.127 (-0.142 to -0.111) ★ + | -0.065 (-0.098 to -0.034) ★ | -0.105 (-0.137 to -0.075) ★ |
|  | HF | 0.003 (-0.007 to 0.009) | 0.015 (0.011 to 0.020) ★ | 0.023 (0.006 to 0.041) ★ | -0.002 (-0.022 to 0.017) |
|  | IS | 0.002 (-0.006 to 0.010) | -0.010 (-0.022 to -0.002) ★ | -0.005 (-0.022 to 0.011) | -0.003 (-0.026 to 0.019) |
|  | PCI/CABG | -0.072 (-0.095 to -0.057) ★ + | -0.089 (-0.094 to -0.080) ★ + | -0.062 (-0.097 to -0.030) ★ | -0.091 (-0.120 to -0.064) ★ |
|  | death | 0.058 (0.050 to 0.064) ★ + | 0.077 (0.075 to 0.078) ★ + | 0.043 (0.029 to 0.058) ★ | 0.071 (0.056 to 0.085) ★ |
|  | ACS/HF/IS | -0.001 (-0.003 to 0.001) | -0.020 (-0.022 to -0.018) ★ + | -0.006 (-0.018 to 0.007) | -0.018 (-0.032 to -0.004) ★ |
|  | ACS/HF/IS/death | 0.023 (0.017 to 0.028) ★ + | 0.017 (0.015 to 0.018) ★ + | 0.021 (0.011 to 0.031) ★ | 0.021 (0.010 to 0.032) ★ |
| **A:**<br>HEM<br>AGE<br>HX<br>**B:**<br>AGE<br>HX | ACS | 0.006 (-0.009 to 0.016) | -0.006 (-0.016 to 0.007) | 0.011 (-0.004 to 0.026) | -0.002 (-0.009 to 0.005) |
|  | HF | 0.037 (0.034 to 0.041) ★ + | 0.067 (0.066 to 0.070) ★ + | 0.049 (0.037 to 0.062) ★ | 0.042 (0.029 to 0.056) ★ |
|  | IS | 0.020 (0.012 to 0.026) ★ | 0.017 (0.009 to 0.024) ★ | 0.006 (-0.008 to 0.020) | 0.011 (-0.007 to 0.028) |
|  | PCI/CABG | 0.003 (-0.008 to 0.010) | 0.002 (-0.005 to 0.005) | 0.012 (-0.001 to 0.026) | -0.003 (-0.013 to 0.006) |
|  | death | 0.065 (0.053 to 0.070) ★ + | 0.080 (0.078 to 0.081) ★ + | 0.050 (0.038 to 0.064) ★ | 0.072 (0.059 to 0.086) ★ |
|  | ACS/HF/IS | 0.029 (0.026 to 0.030) ★ + | 0.029 (0.026 to 0.030) ★ + | 0.024 (0.016 to 0.033) ★ | 0.020 (0.013 to 0.028) ★ |
|  | ACS/HF/IS/death | 0.045 (0.044 to 0.046) ★ + | 0.046 (0.045 to 0.047) ★ + | 0.040 (0.032 to 0.048) ★ | 0.041 (0.033 to 0.049) ★ |
| **A:**<br>HEM<br>AGE<br>HX<br>**B:**<br>HEM<br>AGE | ACS | 0.079 (0.061 to 0.115) ★ + | 0.121 (0.105 to 0.145) ★ + | 0.075 (0.044 to 0.107) ★ | 0.103 (0.074 to 0.134) ★ |
|  | HF | 0.034 (0.030 to 0.045) ★ + | 0.052 (0.049 to 0.057) ★ + | 0.026 (0.016 to 0.037) ★ | 0.044 (0.034 to 0.054) ★ |
|  | IS | 0.018 (0.014 to 0.024) ★ + | 0.026 (0.018 to 0.033) ★ + | 0.011 (0.005 to 0.018) ★ | 0.014 (0.002 to 0.027) ★ |
|  | PCI/CABG | 0.075 (0.061 to 0.095) ★ + | 0.091 (0.084 to 0.097) ★ + | 0.074 (0.047 to 0.105) ★ | 0.088 (0.062 to 0.115) ★ |
|  | death | 0.007 (-0.005 to 0.019) | 0.004 (0.003 to 0.005) ★ | 0.007 (0.002 to 0.011) ★ | 0.002 (-0.002 to 0.005) |
|  | ACS/HF/IS | 0.030 (0.028 to 0.031) ★ + | 0.049 (0.046 to 0.051) ★ + | 0.030 (0.022 to 0.038) ★ | 0.038 (0.030 to 0.048) ★ |
|  | ACS/HF/IS/death | 0.022 (0.017 to 0.028) ★ + | 0.030 (0.028 to 0.031) ★ + | 0.019 (0.014 to 0.025) ★ | 0.020 (0.014 to 0.025) ★ |

Mean and 95% confidence interval of the statistic $\Delta_C = C_A - C_B$ , where $C_A$ is the C-index of an outcome modeled with predictor set A, and $C_B$ is the C-index for an outcome modeled with predictor set B. An asterisk ( ★ ) denotes that $\Delta_C$ is significantly different from zero. A plus sign ( + ) next to a derivation-set result denotes that that $\Delta_C$ is also significantly different from zero in the validation set, and the effect is in the same direction as in the derivation dataset, i.e. the result in the derivation set is corroborated by the result in the validation set). There were no cases where a validation-set result was significant, but in the opposite direction of a significant derivation-set result.

The addition of hematologic predictors to the AGE models improved (made smaller) Brier scores for almost all outcomes in the internal test set, for both men and women (**Supplementary Table 4**). Out of 14 comparisons for these two predictor sets, 11 showed a statistically significant change in Brier score on the internal test set. Of those 11, 10 also showed a corroborating statistically significant change on the external validation set. Only the ACS models and the female IS model and did not show a statistically significant change.

#### Incorporating disease history: AGE-HX vs AGE

We hypothesized that history for some diseases would be available in most EHRs, albeit with varying degrees of completeness, and so may be considered for a scalable risk-assessment strategy. Adding disease histories to the AGE predictor set (i.e., making the AGE-HX predictor set) resulted in superior discrimination: AGE-HX models had higher C-index compared to AGE models for all outcomes, for both sexes, and in both derivation and validation cohort (**Table 4**). On the internal test set, increases in C-index were in the range 0.026 to 0.126. On the external validation set, increases in C-index were in the range 0.020 to 0.108. AGE-HX models also had better (lower) Brier scores than the AGE models (**Supplementary Table 4**). Out of 14 comparisons on the internal test set, all 14 showed statistically significant improvement, and all 14 improvements were corroborated by statistically significant improvements in the external validation set.

#### Substitution of hematology for history: AGE-HEM vs AGE-HX

To address the scenario where these highly predictive diagnostic codes are unavailable (e.g., a population-health program initiated with the help of a diagnostic lab provider), we compared the utility of the AGE-HEM models to that of the AGE-HX models. Equivalence between the two predictor sets depended on the outcome considered. For HF, IS, and the composite outcomes, the drop in C-index associated with using AGE-HEM instead of AGE-HX was < 0.03, for men and women, on both internal and external cohorts (**Table 4**). For modeling death, AGE-HEM was in fact superior to AGE-HX by as much as 0.077. For ACS and PCI/CABG, the drop in C-index was between 0.062 and 0.127 and was significant in both sexes and datasets. Calibration showed minimal difference, though for all outcomes except death and ACS/HF/IS/death, using AGE-HEM instead of AGE-HX produced minimally inferior Brier scores (**Supplementary Table 4**), with the HF model showing the lowest magnitude change in performance (increase of 0.003 on the external dataset).

#### Incremental value of hematologic predictors to age + disease history: HEM-AGE-HX vs AGE-HX

As was the case when we added hematologic predictors to the AGE models, we found that adding hematologic predictors to the AGE-HX models (thus making the HEM-AGE-HX models) often yielded significantly higher C-indices than did the AGE-HX predictor set, both for men and for women (**Table 4**). For example, C-index for the male HF model was higher by 0.067 under the larger predictor set, and C-index for the male death model was higher by 0.080 under the larger predictor set. The incremental effect of hematologic predictors was usually smaller than the difference between HEM-AGE and AGE, and so effects were less often significant in both validation and derivation datasets. Models for HF, death, and the composite outcomes showed statistically significant improvements in both cohorts for both sexes. Improvements in Brier score followed almost the same pattern (**Supplementary Table 4**). On the validation cohort, the HEM-AGE-HX models demonstrated C-indices between 0.75 and 0.85 (**Table 3**).

#### Calibration curves

We examined calibration curves for the HEM-AGE-HX model for all outcomes (**Figure 1)**, as measured on the validation cohort. For females, risk is calibrated well for the all but the highest-risk decile. Over-prediction of risk for high-risk patients is more pronounced among the male models than the female models, consistent with higher Brier scores for male models.

**Figure 1.**
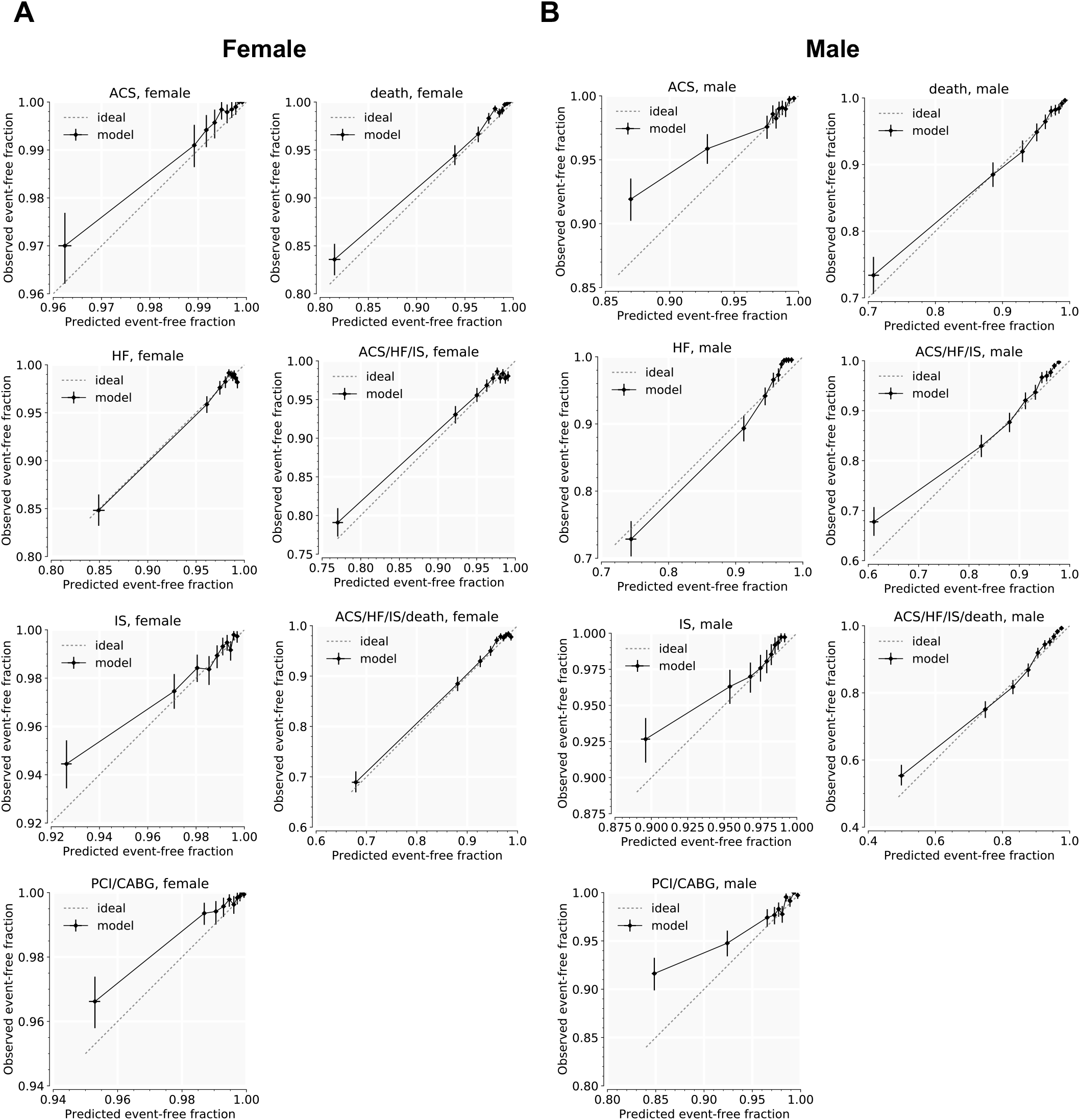
Calibration curves for HEM-AGE-HX models on validation cohort. Calibration curves for the seven modeled outcomes, on female (**A**) and male (**B**) in the validation cohort. Predictor set is HEM-AGE-HX, which includes hematology and age and disease history, and interactions with age. Predicted risk is compared with observed outcomes at 3 years.

#### Survival by predicted-risk quartile

Discrimination was assessed visually using Kaplan-Meier curves for the seven outcomes, as modeled with the HEM-AGE-HX predictor set and measured on the external validation cohort (**Figure 2)**. In each subplot, the cohort is split into quartiles, according to the predicted relative risk of the outcome for each patient.

**Figure 2.**
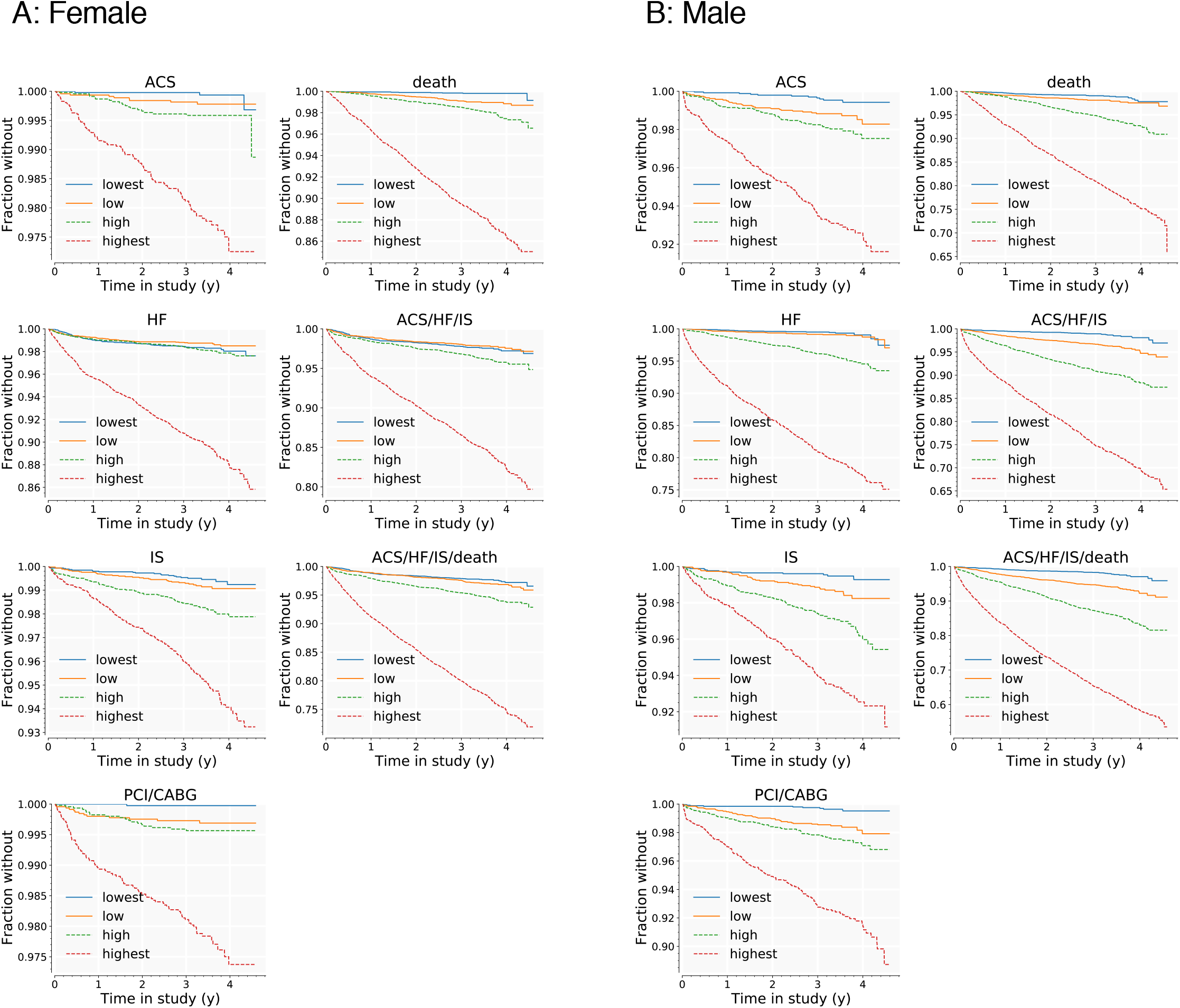
Kaplan-Meier curves for HEM-AGE-HX models, by quartiles of predicted risk in validation cohort. Curves are shown for all seven outcomes in the validation cohort, females (**A**) and males (**B**). For each outcome, the cohort is split into quartiles, according to the risk model developed for that outcome on the derivation cohort. Lowest-risk quartile is shown in blue; highest-risk quartile is shown in red.

For all seven outcomes among men, the risk groups predicted by the models maintain proper rank by observed risk over the whole observation period, at least for the highest risk 3 quartiles. For the HF model, survival curves for the low-risk and lowest-risk quartiles intersect at a follow-up time of around 4 years, which can indicate poor calibration of this model for the healthiest patients, though given the low event rate after 4 years (< 10/month for the entire male cohort), this is also readily attributable to noise in the Kaplan-Meier estimator.

Performance for the female-cohort models is similar: survival curves for the riskiest three quartiles are properly ordered over the entire observation period, but the low-risk and lowest-risk curves are sometimes inverted for the HF model and for the two composite models. The only exception is the PCI/CABG model, for which the middle two risk groups are inverted at t < 2 years.

### Parameter estimates

Proportional-hazards coefficients in the HEM-AGE-HX models were examined for the three outcomes (composites not included) where the HEM-AGE-HX predictor set had better discrimination than AGE-HX on the internal test set (**Table 5**). The *L*_*1*_ penalization selects approximately half of the ten hematologic indices for each of the final models. Among the highly correlated set {HCT, HGB, MCH, MCHC, MCV, RBC}, usually more than half are eliminated, with the most commonly retained being RBC and HGB. Two out of the ten hematologic indices appear in all six models in Table 8: platelet count (PLT) and red cell distribution width (RDW). White cell count (WBC) and red cell count (RBC) are retained in five out of six models. As expected, the disease-history terms and the age terms are extremely strong factors in most HEM-AGE-HX models.

**Table 5.** HEM-AGE-HX parameter estimates for selected outcomes.

| Predictor | HF Hospitalization |  |  |  | Ischemic Stroke |  |  |  | Death |  |  |  |
| --- | --- | --- | --- | --- | --- | --- | --- | --- | --- | --- | --- | --- |
|  | Female |  | Male |  | Female |  | Male |  | Female |  | Male |  |
| | $\beta$ | $p$ | $\beta$ | $p$ | $\beta$ | $p$ | $\beta$ | $p$ | $\beta$ | $p$ | $\beta$ | $p$ |
| HCT | 0 | 1 | 0 | 0.999 | 0 | 1 | 0 | 1 | 0 | 0.999 | 0 | 0.999 |
| HGB | -0.01 | 0.945 | 0 | 0.999 | -0.078 | 0.534 | -0.266 | 0.001 | -0.468 | 0.029 | -0.172 | 0.021 |
| MCH | 0 | 1 | 0 | 0.999 | 0 | 1 | -0.002 | 0.983 | 0 | 0.999 | 0 | 0.999 |
| MCHC | 0 | 1 | 0 | 0.999 | 0 | 1 | -0.014 | 0.875 | 0.14 | 0.1 | 0 | 0.999 |
| MCV | 0.135 | 0.154 | 0 | 0.999 | 0 | 1 | 0 | 1 | 0.318 | 0.008 | 0 | 0.999 |
| MPV | -0.046 | 0.299 | 0 | 0.999 | 0 | 1 | 0 | 1 | -0.092 | 0.034 | -0.005 | 0.892 |
| PLT | -0.194 | 0.002 | -0.049 | 0.157 | -0.089 | 0.288 | 0.002 | 0.972 | -0.105 | 0.009 | -0.242 | 0 |
| RBC | -0.053 | 0.704 | -0.334 | 0 | -0.211 | 0.061 | 0 | 1 | -0.003 | 0.989 | -0.357 | 0 |
| RDW | 0.376 | 0 | 0.261 | 0 | 0.207 | 0.007 | 0.014 | 0.836 | 0.332 | 0 | 0.286 | 0 |
| WBC | 0.043 | 0.321 | 0 | 1 | 0.051 | 0.166 | 0.082 | 0.001 | 0.081 | 0.006 | 0.02 | 0.354 |
| age | 0.473 | 0 | 0.072 | 0.206 | 0.714 | 0 | 0.514 | 0 | 0.799 | 0 | 0.519 | 0 |
| Hx_CAD | 0.548 | 0.005 | 0.249 | 0.034 | 0.408 | 0.192 | 0 | 1 | 0.479 | 0.024 | 0.166 | 0.237 |
| Hx_HF | 2.149 | 0 | 1.576 | 0 | 0.578 | 0.066 | 0.207 | 0.221 | 0.792 | 0 | 0.539 | 0 |
| Hx_stroke | 0 | 1 | 0.019 | 0.904 | 1.281 | 0 | 1.64 | 0 | 0 | 0.999 | 0.024 | 0.866 |
| HCT × age | 0 | 1 | 0 | 0.999 | 0 | 1 | 0 | 1 | 0 | 0.999 | 0.053 | 0.163 |
| HGB × age | -0.056 | 0.426 | 0 | 1 | 0 | 1 | -0.088 | 0.168 | 0 | 0.999 | 0 | 0.999 |
| MCHC × age | 0 | 1 | -0.016 | 0.647 | 0.023 | 0.714 | 0 | 1 | 0 | 0.999 | 0 | 0.999 |
| MCH × age | 0 | 1 | 0 | 0.999 | 0 | 1 | 0 | 1 | -0.001 | 0.991 | 0 | 1 |
| MCV × age | 0 | 1 | 0 | 1 | -0.059 | 0.358 | 0.097 | 0.112 | -0.034 | 0.74 | 0 | 0.999 |
| MPV × age | 0.033 | 0.288 | 0 | 1 | 0.027 | 0.533 | -0.001 | 0.978 | 0.06 | 0.042 | 0.022 | 0.481 |
| PLT × age | 0.005 | 0.927 | 0 | 0.999 | -0.006 | 0.939 | 0.066 | 0.269 | 0 | 1 | 0.101 | 0.01 |
| RBC × age | -0.024 | 0.707 | 0 | 0.999 | 0 | 1 | 0 | 1 | 0.068 | 0.135 | 0 | 0.999 |
| RDW × age | -0.04 | 0.302 | 0 | 0.999 | -0.011 | 0.858 | 0 | 1 | -0.049 | 0.181 | -0.008 | 0.791 |
| WBC × age | -0.02 | 0.51 | 0 | 1 | 0 | 1 | -0.034 | 0.489 | -0.014 | 0.541 | 0 | 1 |
| age × age | 0.121 | 0.005 | 0 | 1 | 0 | 1 | 0.026 | 0.666 | 0.085 | 0.043 | 0.038 | 0.345 |
| Hx_CAD × age | -0.007 | 0.96 | 0.093 | 0.341 | -0.043 | 0.851 | -0.104 | 0.398 | -0.155 | 0.292 | 0.061 | 0.607 |
| Hx_HF × age | -0.623 | 0 | 0 | 0.999 | -0.329 | 0.159 | 0 | 1 | -0.22 | 0.113 | 0.056 | 0.616 |
| Hx_stroke × age | 0.236 | 0.042 | 0 | 0.999 | -0.026 | 0.911 | -0.482 | 0.005 | 0.127 | 0.256 | 0 | 1 |

### Random Survival Forest

Discrimination of the Cox proportional hazards models was comparable to the discrimination of the random survival forests (**Supplementary Table 6**). Many of the optimized RSF models had slightly better discrimination than the Cox proportional hazards models, with advantages for RSF ranging from −0.009 to 0.050. The biggest advantage for RSF came when using the HEM predictor set, which does not include any explicit interaction terms, and which therefore might be harder for a linear model to learn from than for a tree-based model. Improvement was less apparent for the HEM-AGE-HX models.

## Discussion

The objective of our work is fundamentally different from that of most efforts focused on training risk models: we sought to develop risk models for cardiovascular events based entirely on low-cost, widely available, well-calibrated^20^, and objective input features. Whereas most risk models have emphasized the utility of a diverse set of inputs, which will typically require providers to order specific laboratory tests, document an expansive set of diagnoses, and record candid responses of patients regarding such attributes as smoking status^18^, our approach deliberately attempts to minimize the burden on providers or dependence on costly elements of data collection. Our rationale is based in part on our own experience with EHR-based risk assessment: for example, the percentage of patients in our cohorts for which one can compute the Pooled-Cohort Equation risk score from structured data within the MGB-EDW is <30% (data not shown). Even when the relevant tests may have been ordered, they may not be available as structured data in the EHR, if, for example, they were collected outside of the system and were only available as scanned documents. Furthermore, most risk models focus on primary prevention – whereas our focus was on a broader patient population with anticipated high resource utilization.

We have used hematologic predictors based on an assumption that many contributors to chronic diseases such as cardiovascular disease are systemic and may be reflected across multiple tissue types. This hypothesis is supported by a large amount of prior data, both experimental and observational, including the ability to estimate cardiovascular outcomes from digital retinograms^21^, the association of somatic mutations in leukocytes with coronary artery disease and heart failure risk^6,22^, the association of a complete-blood count-based score with mortality^23–27^, and the modulation of cardiovascular disease through leukocyte-restricted gene knockouts^28,29^.

Others have shown association of hematologic indices with all-cause mortality and cardiovascular events, including a focus on red-cell distribution width^23,30–34^ and specific leukocyte subpopulations such as the neutrophil-to-lymphocyte ratio^35^. The utility of RDW for predicting outcomes may be related to its association with anemias of chronic disease or with clonal hematopoiesis of indeterminate potential^6^, which appear to predict similar patterns of events. In both cases, the biomarkers may primarily be providing a readout for underlying systemic pathway abnormalities, with the hematopoietic lineage representing a shared upstream pathway abnormality or an index of prior exposures.

Although our work demonstrated success primarily for heart failure, major adverse cardiovascular events, all-cause mortality, and to a lesser extent ischemic stroke, we had less success in predicting incident acute coronary syndromes beyond what is possible with age and prior diagnoses. Nonetheless, a model using hematologic indices alone was able to modestly predict incident acute coronary syndromes (C-index = 0.62 [95% CI 0.57 to 0.67] and 0.60 [95% CI 0.56 to 0.63] for women and men, respectively, on validation dataset, comparable to polygenic risk scores^2^) suggesting that some signal exists, but there is likely redundancy with age and existing CVD. Nonetheless, a strategy combining age and hematologic predictors may still be useful in situations where structured diagnoses are not available or as complete as found within the MGB-EDW.

The primary limitation of our work, which is also a strength, is a deliberate use of restricted numbers of inputs to the risk models. Our patient population may also exhibit some selection bias that could limit generalizability to other settings – though in such cases, our approach combining widely available inputs with automated adjudication should enable rescaling or retraining. We have also not looked at out how these measures of risk progress through time and in response to therapy in individual patients. Given the limited performance benefit of random survival forests for predicting outcomes with these same inputs, we chose not to explore other algorithms, though it is possible some would show superior performance.

There is no shortage of risk models available for cardiovascular outcomes – and the challenge is motivating their use. We suspect that models such as the ones described here will primarily be attractive at an institutional or payer level, where better estimation of risk for a very large percentage of a population has clear economic value. We are not aware of any comparable effort to deliberately push the limits of models with minimal low-cost input features. We see our approach as complementary to other models which typically rely on individual or provider-entered data and have focused on patients not currently taking statins and free of cardiovascular disease^17,18,36^. These approaches also lend themselves to the creation of continuously updating risk trajectories where the rates of change may have additional predictive utility. Our goal is to maximize the number of patients for which a system-level estimation of risk is feasible, with minimal disruption to the existing workflow of providers, a place where we see model deployment having the greatest impact.

## Data Availability

The data that support the findings of this study are available on request from the corresponding author R.C.D. upon approval of the data sharing committees of the respective institutions. The data are not publicly available due to the presence of information that could compromise research participant privacy.

## Supplementary Figures

**Supplementary Figure 1.**
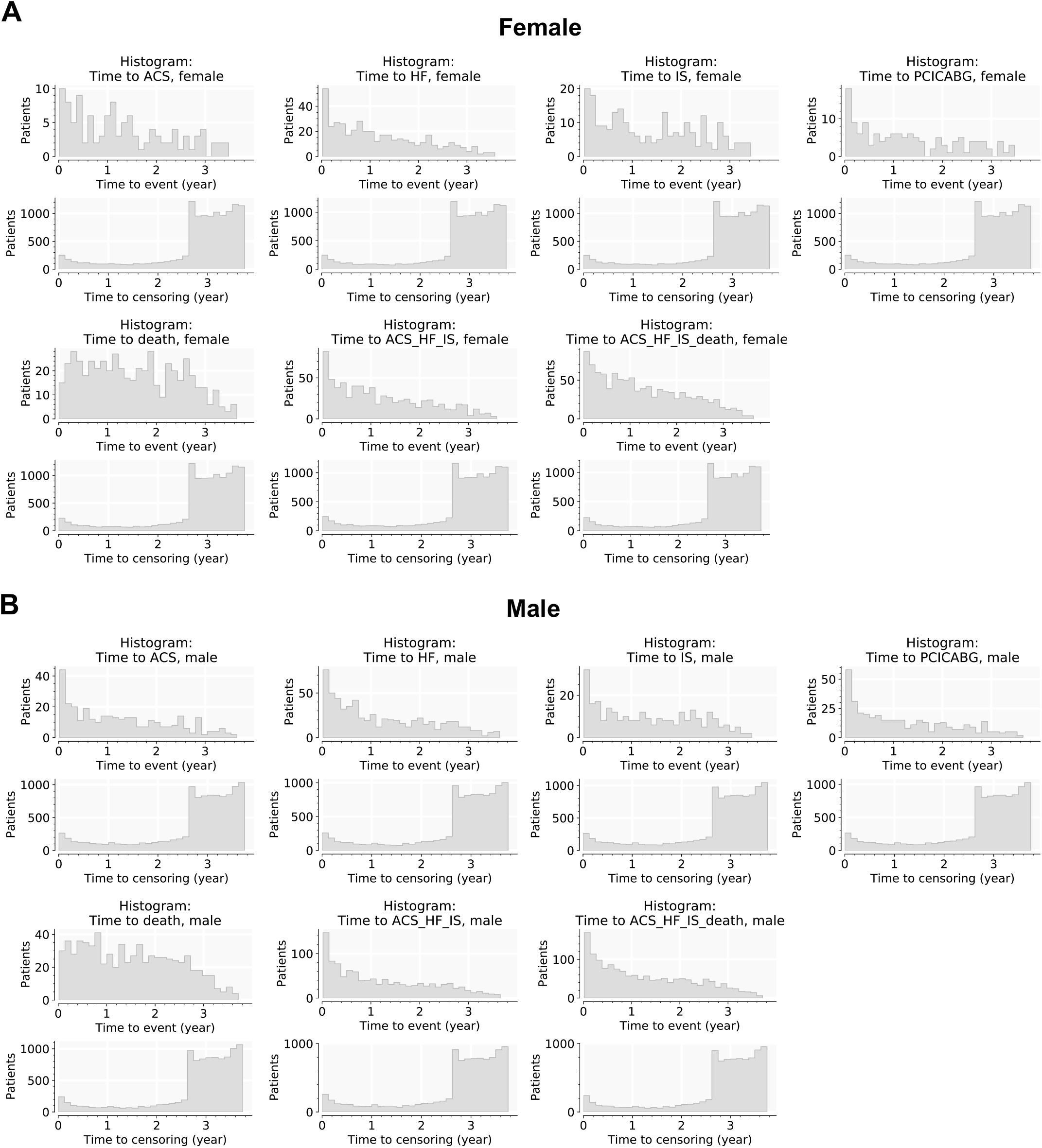
Distribution of time to events in derivation cohort. Time to 1^st^ outcome, also time to right-censoring, for patients without outcomes in the observation period. Results broken down by the seven outcomes for female (**A**) and male (**B**) in this study. Derivation cohort.

**Supplementary Figure 2.**
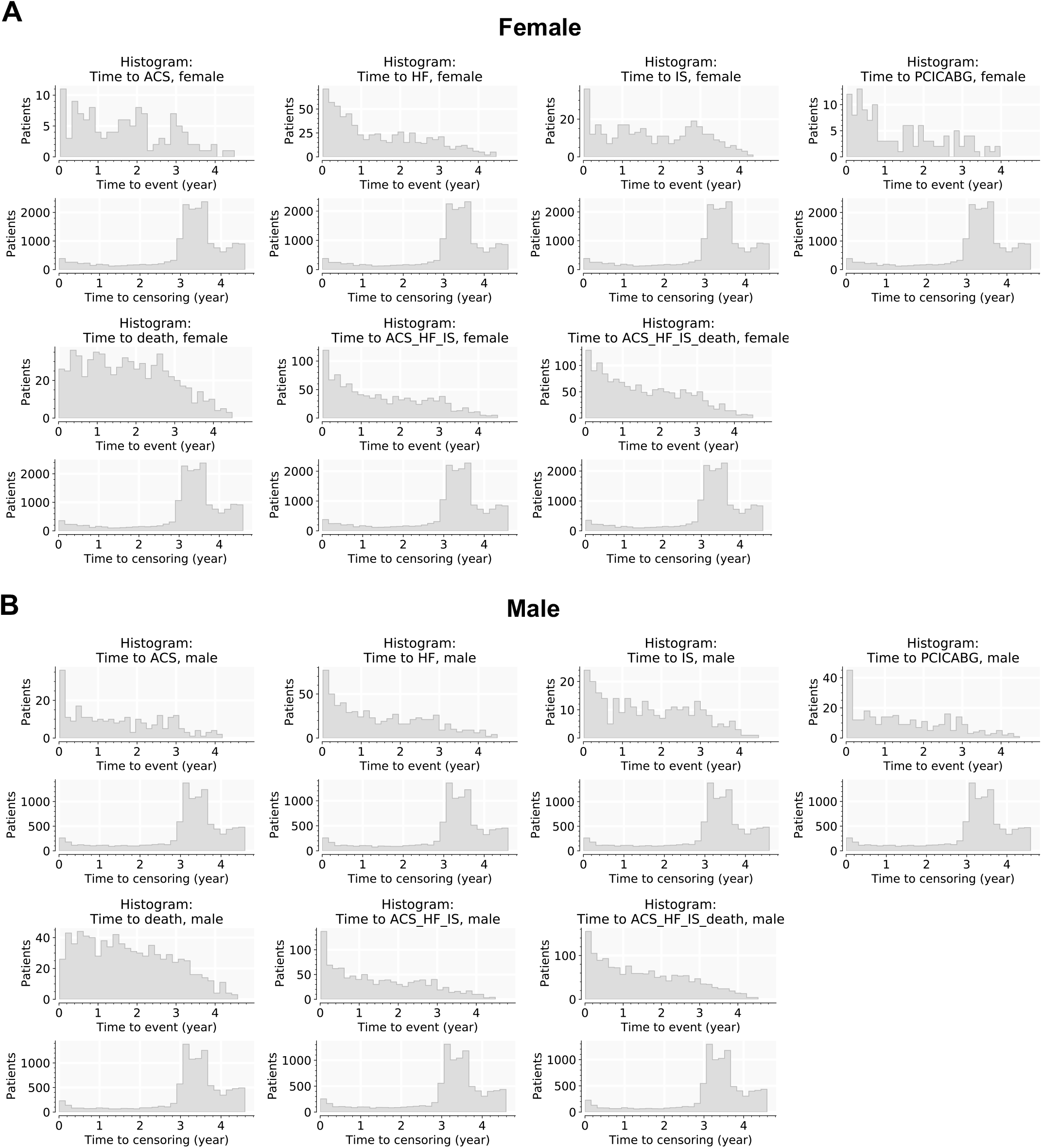
Distribution of time to events in validation cohort. Time to 1^st^ outcome, also time to right-censoring, for patients without outcomes in the observation period. Results broken down by the seven outcomes for female (**A**) and male (**B**) in this study.

**Supplementary Figure 3A:**
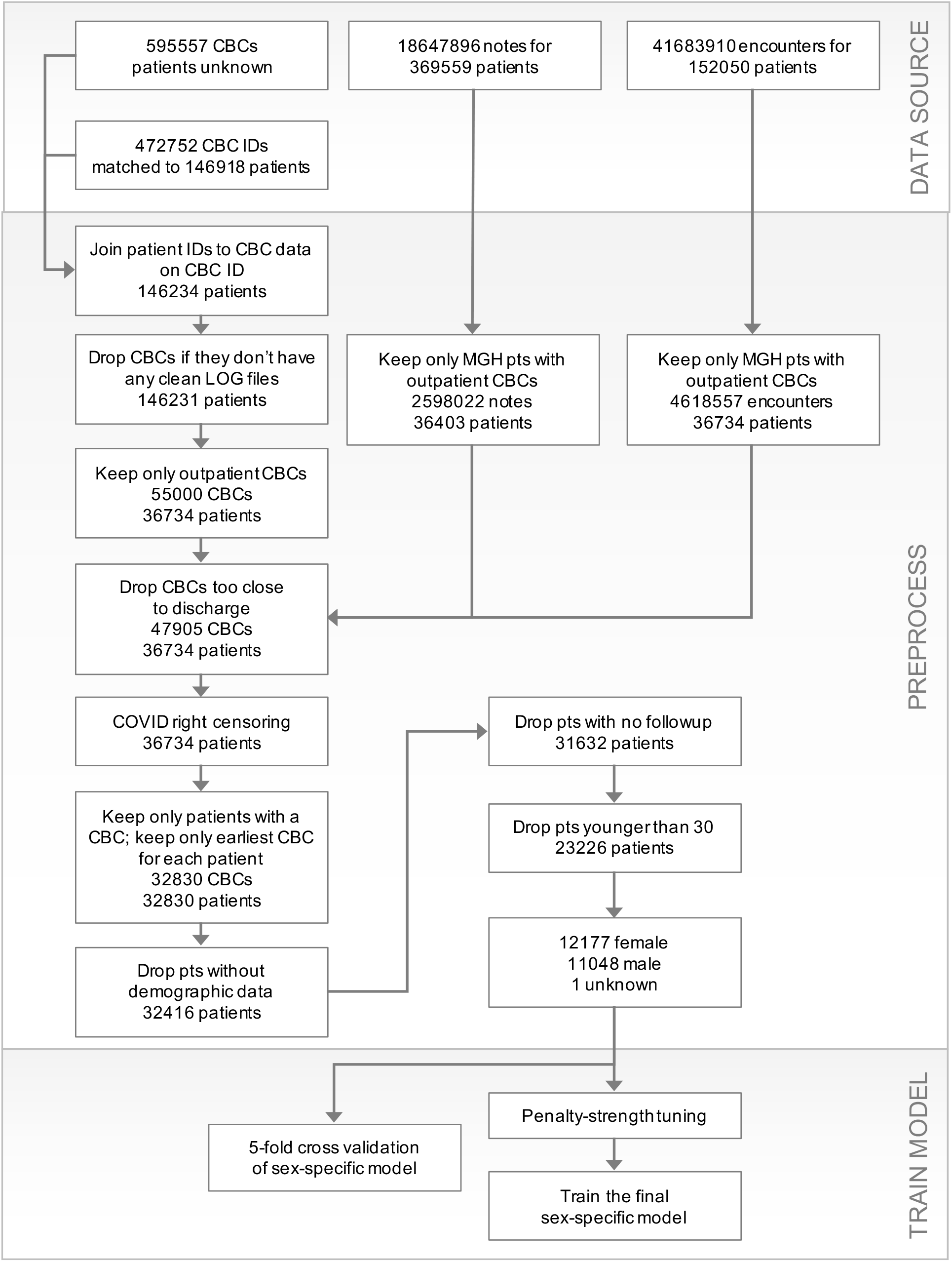
Flow diagram for modeling CVD events in derivation cohort. MGH patients and their data, used to model time-to-event for ACS, HF hospitalization, stroke, PCI/CABG

**Supplementary Figure 3B:**
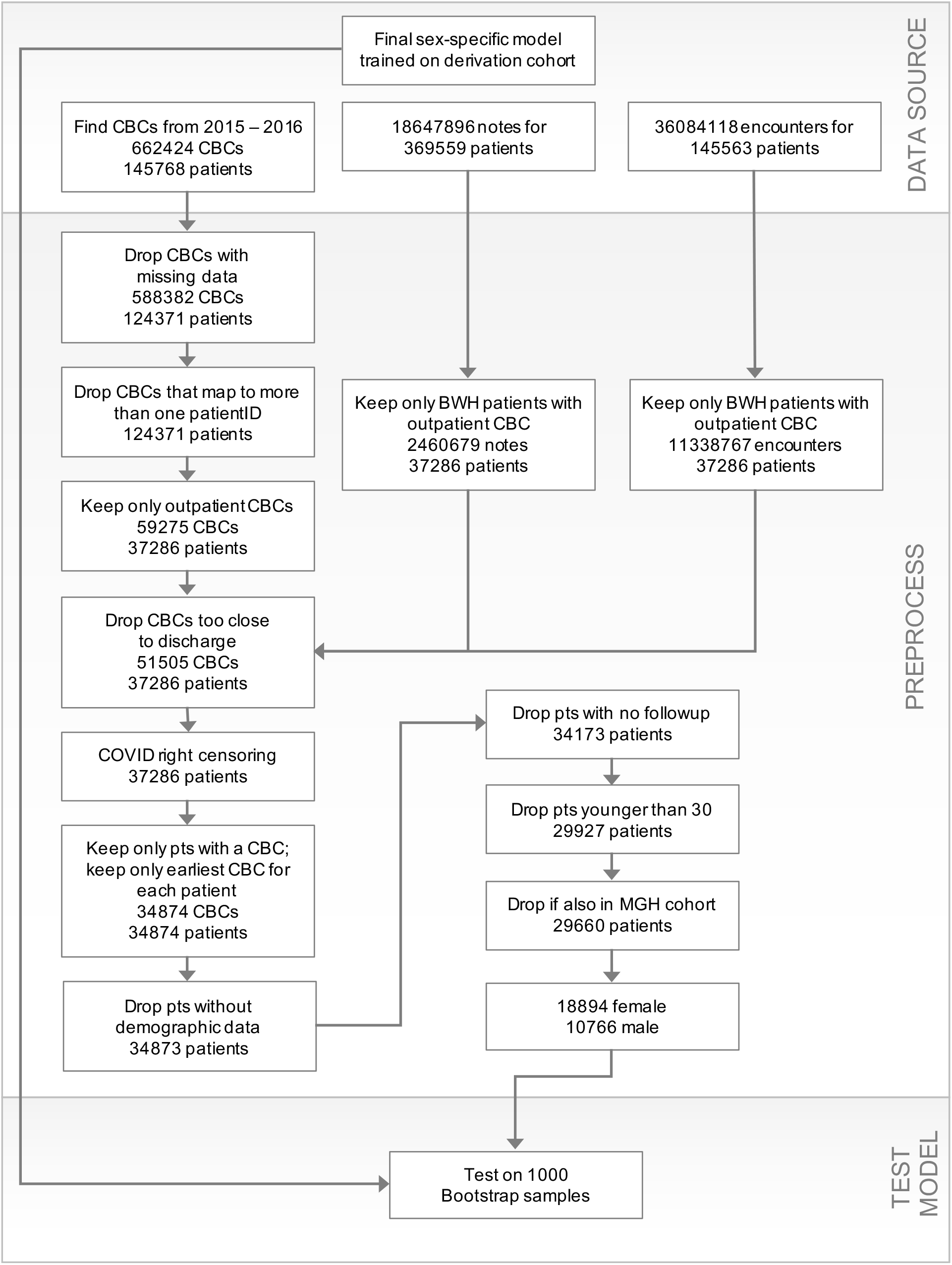
Flow diagram for modeling CVD events in validation cohort. BWH patients and their data, used to model time-to-event for ACS, HF hospitalization, stroke, PCI/CABG

**Supplementary Figure 4A:**
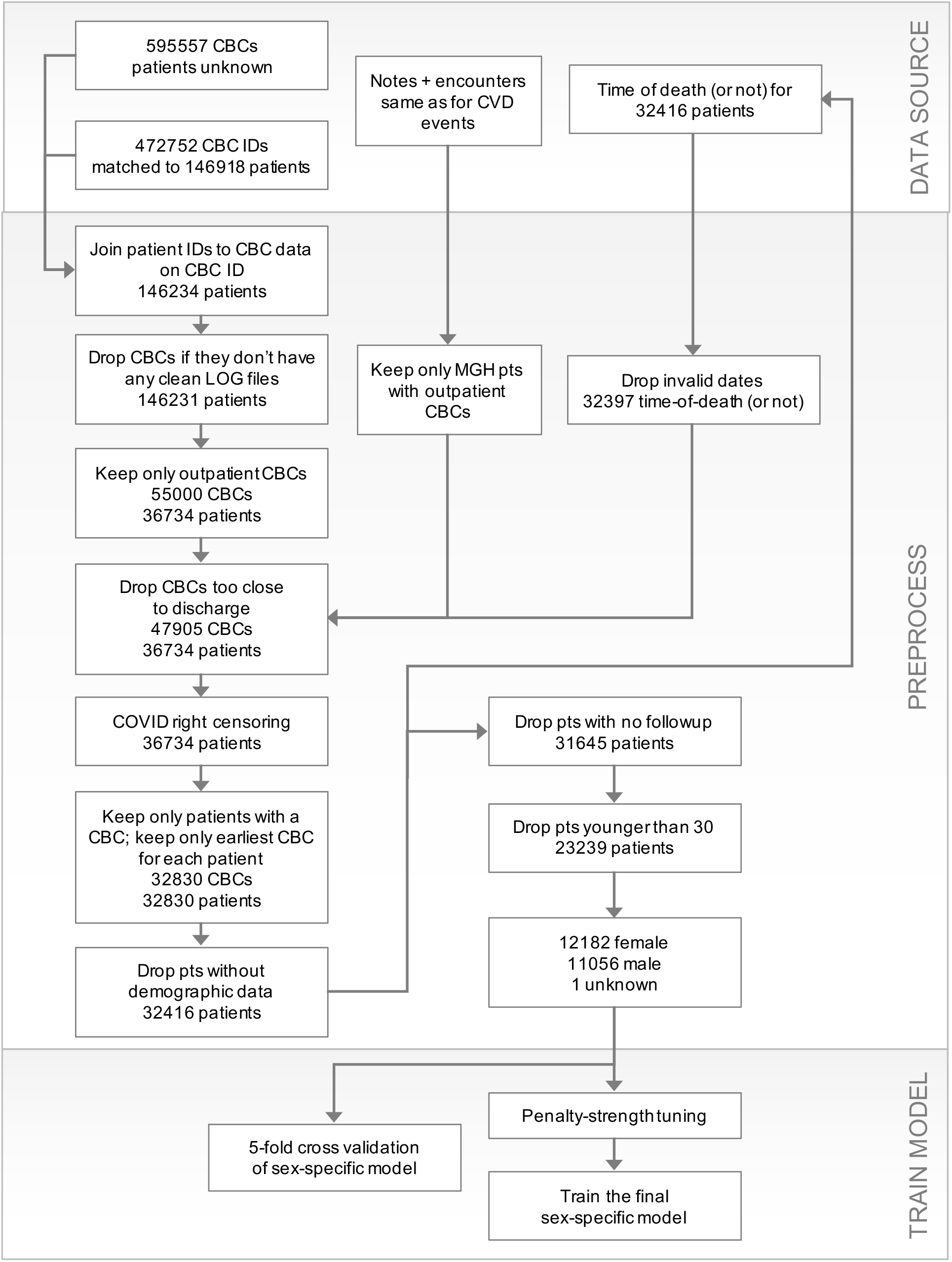
Flow diagram for modeling death in derivation cohort. MGH patients and their data, used to model time-to-event for death

**Supplementary Figure 4B:**
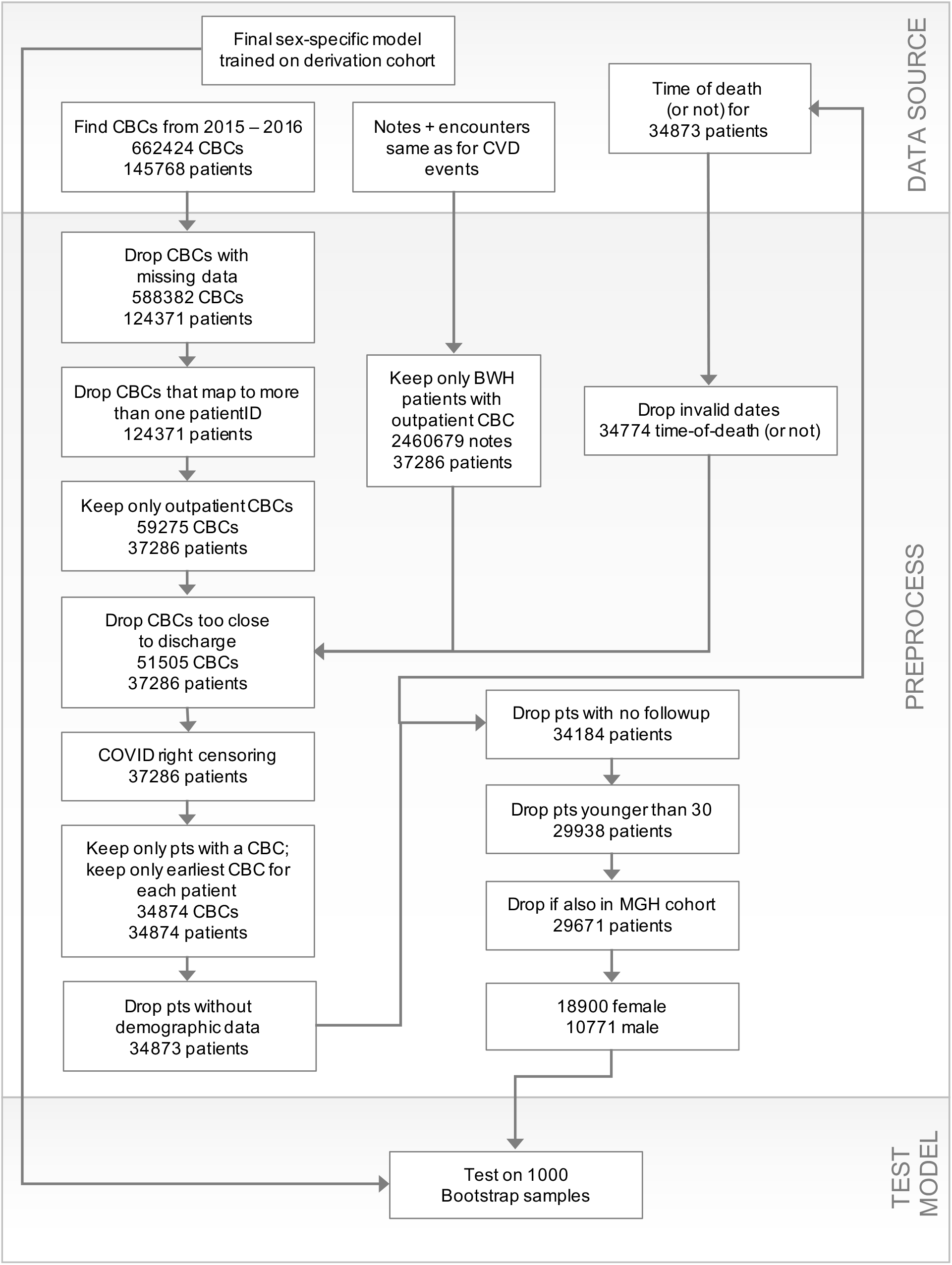
Flow diagram for modeling death in validation cohort. BWH patients and their data, used to model time-to-event for death

## Supplementary Tables

**Supplementary Table 1A.** Performance of artificial-intelligence models for CVD-event adjudication. Performance of classifier algorithms used to adjudicate outcomes for our study. Classification thresholds were chosen to maximize F1 score.

| Event | Sensitivity | Specificity | Precision |
| --- | --- | --- | --- |
| ACS | 0.76 | 0.97 | 0.59 |
| HF | 0.94 | 0.91 | 0.58 |
| IS | 0.80 | 0.97 | 0.64 |
| PCI/CABG | 0.98 | 0.99 | 0.90 |

**Supplementary Table 1B.** Performance of ICD codes in predicting history of disease. ICD codes can predict a patient’s history of disease at the time of their entry into the survival study.

| Past History | Sensitivity | Specificity |
| --- | --- | --- |
| ACS | 0.78 | 0.95 |
| HF | 0.68 | 0.95 |
| IS | 0.95 | 0.93 |

**Supplementary Table 2.** Main effects, notation and description. Main effects of the proportional-hazards models referred to in this study. All main effects are unitless in the proportional-hazards model. Except for the three history predictors, all predictors are mean-centered and scaled by their standard deviation, as measured in the derivation cohort. The history variables are binary (0 for no history; 1 for yes history of disease) and are not scaled or centered.

| **Predictor**<br>**Symbol** | **Description of predictor** |
| --- | --- |
| HCT | hematocrit |
| HGB | hemoglobin concentration |
| MCH | mean corpuscular hemoglobin |
| MCHC | mean corpuscular hemoglobin concentration |
| MCV | mean corpuscular volume |
| MPV | mean platelet volume |
| PLT | platelet count |
| RBC | red blood cell count |
| RDW | red cell volume distribution width (C.V.) |
| WBC | white blood cell count |
| age | patient's age at time of entry into survival study |
| Hx_CAD | history of CAD or MI before entry into study |
| Hx_HF | history of HF before entry into study |
| Hx_stroke | history of stroke before entry into study |

**Supplementary Table 3.** Terms used in the five predictor sets for survival models. The elements in each list are the terms, *X*_*i*_, in the linear predictor of the Cox proportional-hazards model.

| Predictor Set | Terms in predictor set |
| --- | --- |
| HEM | HCT, HGB, MCH, MCHC, MCV, MPV, PLT, RBC, RDW, WBC |
| HEM-AGE | HCT, HGB, MCH, MCHC, MCV, MPV, PLT, RBC, RDW, WBC, age,<br>HCT × age, HGB × age, MCHC × age, MCH × age, MCV × age, MPV × age, PLT × age,<br>RBC × age, RDW × age, WBC × age, age × age |
| HEM-AGE-HX | HCT, HGB, MCH, MCHC, MCV, MPV, PLT, RBC, RDW, WBC, age, Hx_CAD, Hx_HF, Hx_stroke,<br>HCT × age, HGB × age, MCHC × age, MCH × age, MCV × age, MPV × age, PLT × age, RBC ×<br>age, RDW × age, WBC × age, age × age, Hx_CAD × age, Hx_HF × age, Hx_stroke × age |
| AGE-HX | age, Hx_CAD, Hx_HF, Hx_stroke, age × age, Hx_CAD × age, Hx_HF × age, Hx_stroke × age |
| AGE | age, age × age, |

**Supplementary Table 4.**
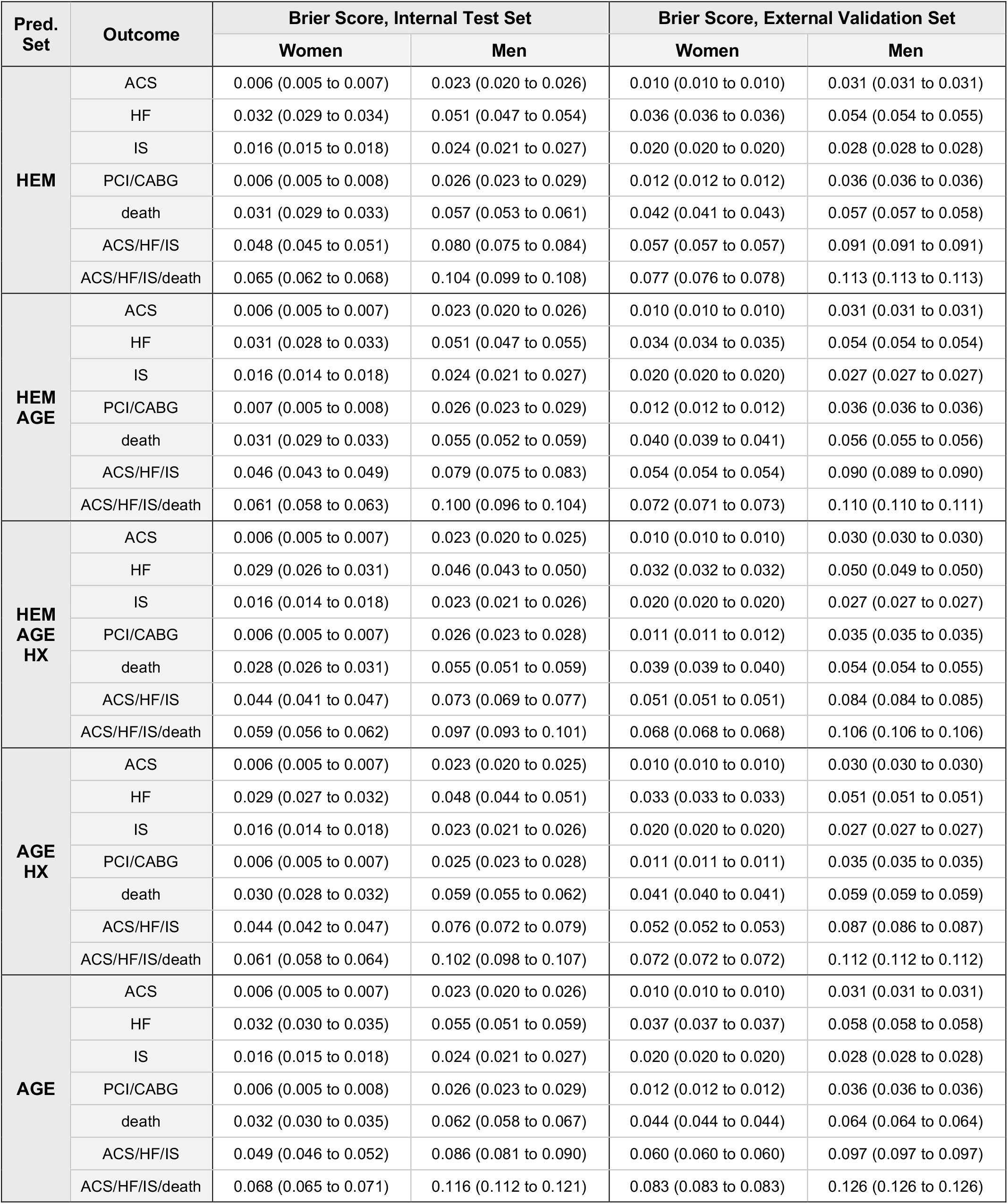
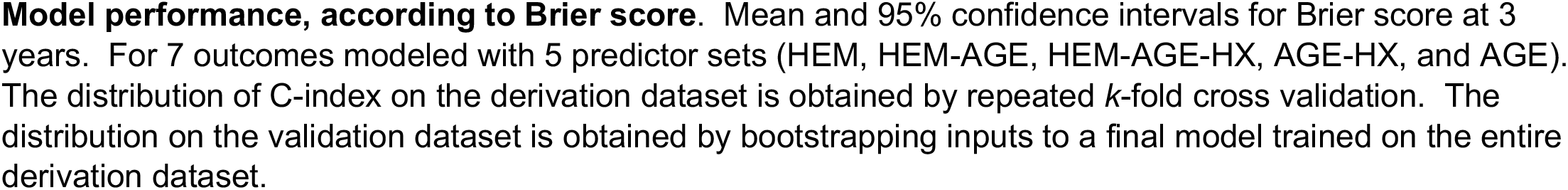
Model performance, according to Brier score at 3 years.

| Pred. Set | Outcome | Brier Score, Internal Test Set |  | Brier Score, External Validation Set |  |
| --- | --- | --- | --- | --- | --- |
|  |  | Women | Men | Women | Men |
| HEM | ACS | 0.006 (0.005 to 0.007) | 0.023 (0.020 to 0.026) | 0.010 (0.010 to 0.010) | 0.031 (0.031 to 0.031) |
|  | HF | 0.032 (0.029 to 0.034) | 0.051 (0.047 to 0.054) | 0.036 (0.036 to 0.036) | 0.054 (0.054 to 0.055) |
|  | IS | 0.016 (0.015 to 0.018) | 0.024 (0.021 to 0.027) | 0.020 (0.020 to 0.020) | 0.028 (0.028 to 0.028) |
|  | PCI/CABG | 0.006 (0.005 to 0.008) | 0.026 (0.023 to 0.029) | 0.012 (0.012 to 0.012) | 0.036 (0.036 to 0.036) |
|  | death | 0.031 (0.029 to 0.033) | 0.057 (0.053 to 0.061) | 0.042 (0.041 to 0.043) | 0.057 (0.057 to 0.058) |
|  | ACS/HF/IS | 0.048 (0.045 to 0.051) | 0.080 (0.075 to 0.084) | 0.057 (0.057 to 0.057) | 0.091 (0.091 to 0.091) |
|  | ACS/HF/IS/death | 0.065 (0.062 to 0.068) | 0.104 (0.099 to 0.108) | 0.077 (0.076 to 0.078) | 0.113 (0.113 to 0.113) |
| HEM AGE | ACS | 0.006 (0.005 to 0.007) | 0.023 (0.020 to 0.026) | 0.010 (0.010 to 0.010) | 0.031 (0.031 to 0.031) |
|  | HF | 0.031 (0.028 to 0.033) | 0.051 (0.047 to 0.055) | 0.034 (0.034 to 0.035) | 0.054 (0.054 to 0.054) |
|  | IS | 0.016 (0.014 to 0.018) | 0.024 (0.021 to 0.027) | 0.020 (0.020 to 0.020) | 0.027 (0.027 to 0.027) |
|  | PCI/CABG | 0.007 (0.005 to 0.008) | 0.026 (0.023 to 0.029) | 0.012 (0.012 to 0.012) | 0.036 (0.036 to 0.036) |
|  | death | 0.031 (0.029 to 0.033) | 0.055 (0.052 to 0.059) | 0.040 (0.039 to 0.041) | 0.056 (0.055 to 0.056) |
|  | ACS/HF/IS | 0.046 (0.043 to 0.049) | 0.079 (0.075 to 0.083) | 0.054 (0.054 to 0.054) | 0.090 (0.089 to 0.090) |
|  | ACS/HF/IS/death | 0.061 (0.058 to 0.063) | 0.100 (0.096 to 0.104) | 0.072 (0.071 to 0.073) | 0.110 (0.110 to 0.111) |
| HEM AGE HX | ACS | 0.006 (0.005 to 0.007) | 0.023 (0.020 to 0.025) | 0.010 (0.010 to 0.010) | 0.030 (0.030 to 0.030) |
|  | HF | 0.029 (0.026 to 0.031) | 0.046 (0.043 to 0.050) | 0.032 (0.032 to 0.032) | 0.050 (0.049 to 0.050) |
|  | IS | 0.016 (0.014 to 0.018) | 0.023 (0.021 to 0.026) | 0.020 (0.020 to 0.020) | 0.027 (0.027 to 0.027) |
|  | PCI/CABG | 0.006 (0.005 to 0.007) | 0.026 (0.023 to 0.028) | 0.011 (0.011 to 0.012) | 0.035 (0.035 to 0.035) |
|  | death | 0.028 (0.026 to 0.031) | 0.055 (0.051 to 0.059) | 0.039 (0.039 to 0.040) | 0.054 (0.054 to 0.055) |
|  | ACS/HF/IS | 0.044 (0.041 to 0.047) | 0.073 (0.069 to 0.077) | 0.051 (0.051 to 0.051) | 0.084 (0.084 to 0.085) |
|  | ACS/HF/IS/death | 0.059 (0.056 to 0.062) | 0.097 (0.093 to 0.101) | 0.068 (0.068 to 0.068) | 0.106 (0.106 to 0.106) |
| AGE HX | ACS | 0.006 (0.005 to 0.007) | 0.023 (0.020 to 0.025) | 0.010 (0.010 to 0.010) | 0.030 (0.030 to 0.030) |
|  | HF | 0.029 (0.027 to 0.032) | 0.048 (0.044 to 0.051) | 0.033 (0.033 to 0.033) | 0.051 (0.051 to 0.051) |
|  | IS | 0.016 (0.014 to 0.018) | 0.023 (0.021 to 0.026) | 0.020 (0.020 to 0.020) | 0.027 (0.027 to 0.027) |
|  | PCI/CABG | 0.006 (0.005 to 0.007) | 0.025 (0.023 to 0.028) | 0.011 (0.011 to 0.011) | 0.035 (0.035 to 0.035) |
|  | death | 0.030 (0.028 to 0.032) | 0.059 (0.055 to 0.062) | 0.041 (0.040 to 0.041) | 0.059 (0.059 to 0.059) |
|  | ACS/HF/IS | 0.044 (0.042 to 0.047) | 0.076 (0.072 to 0.079) | 0.052 (0.052 to 0.053) | 0.087 (0.086 to 0.087) |
|  | ACS/HF/IS/death | 0.061 (0.058 to 0.064) | 0.102 (0.098 to 0.107) | 0.072 (0.072 to 0.072) | 0.112 (0.112 to 0.112) |
| AGE | ACS | 0.006 (0.005 to 0.007) | 0.023 (0.020 to 0.026) | 0.010 (0.010 to 0.010) | 0.031 (0.031 to 0.031) |
|  | HF | 0.032 (0.030 to 0.035) | 0.055 (0.051 to 0.059) | 0.037 (0.037 to 0.037) | 0.058 (0.058 to 0.058) |
|  | IS | 0.016 (0.015 to 0.018) | 0.024 (0.021 to 0.027) | 0.020 (0.020 to 0.020) | 0.028 (0.028 to 0.028) |
|  | PCI/CABG | 0.006 (0.005 to 0.008) | 0.026 (0.023 to 0.029) | 0.012 (0.012 to 0.012) | 0.036 (0.036 to 0.036) |
|  | death | 0.032 (0.030 to 0.035) | 0.062 (0.058 to 0.067) | 0.044 (0.044 to 0.044) | 0.064 (0.064 to 0.064) |
|  | ACS/HF/IS | 0.049 (0.046 to 0.052) | 0.086 (0.081 to 0.090) | 0.060 (0.060 to 0.060) | 0.097 (0.097 to 0.097) |
|  | ACS/HF/IS/death | 0.068 (0.065 to 0.071) | 0.116 (0.112 to 0.121) | 0.083 (0.083 to 0.083) | 0.126 (0.126 to 0.126) |

**Model performance, according to Brier score.** Mean and 95% confidence intervals for Brier score at 3 years. For 7 outcomes modeled with 5 predictor sets (HEM, HEM-AGE, HEM-AGE-HX, AGE-HX, and AGE). The distribution of C-index on the derivation dataset is obtained by repeated *k*-fold cross validation. The distribution on the validation dataset is obtained by bootstrapping inputs to a final model trained on the entire derivation dataset.

**Supplementary Table 5.**
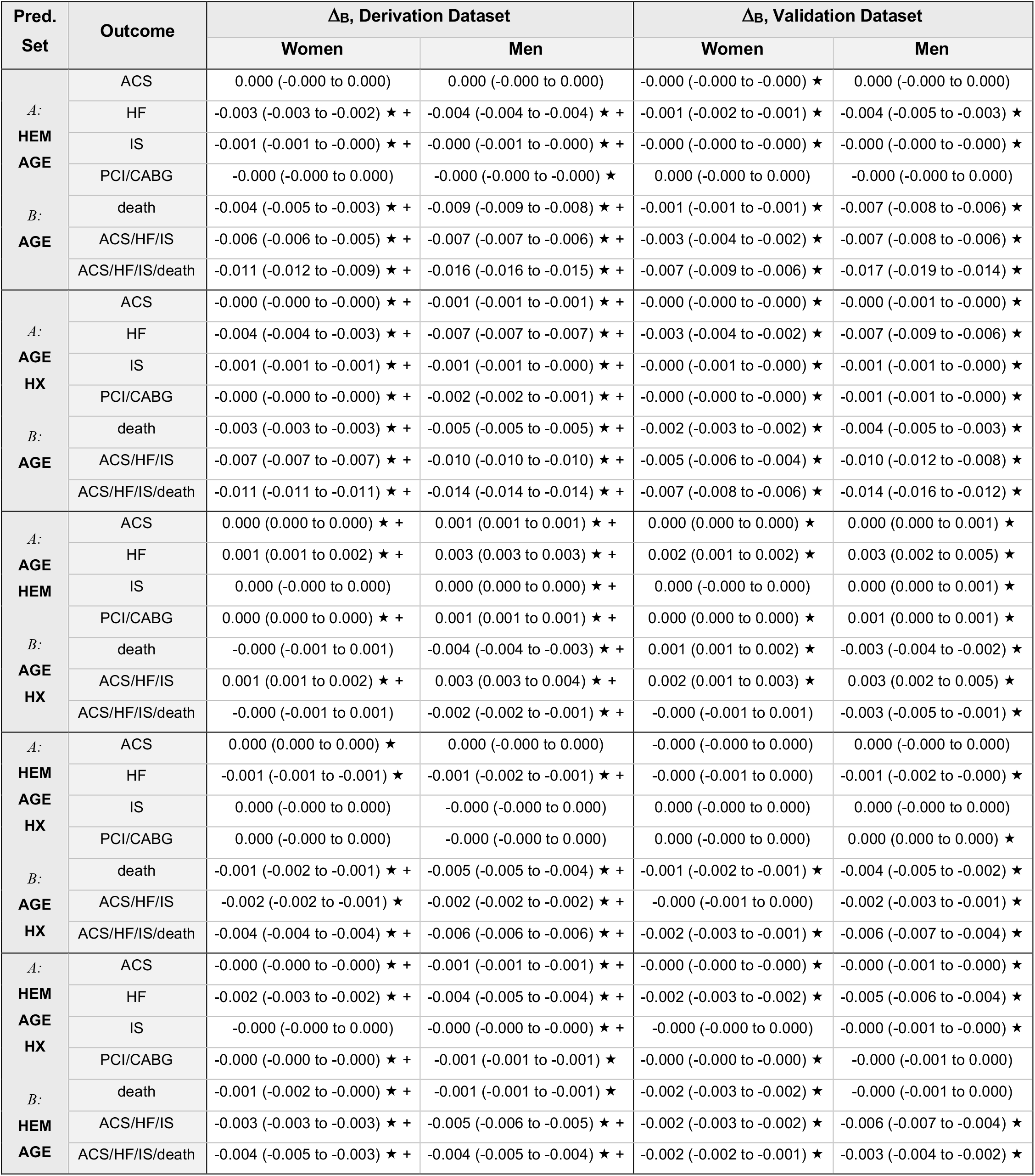
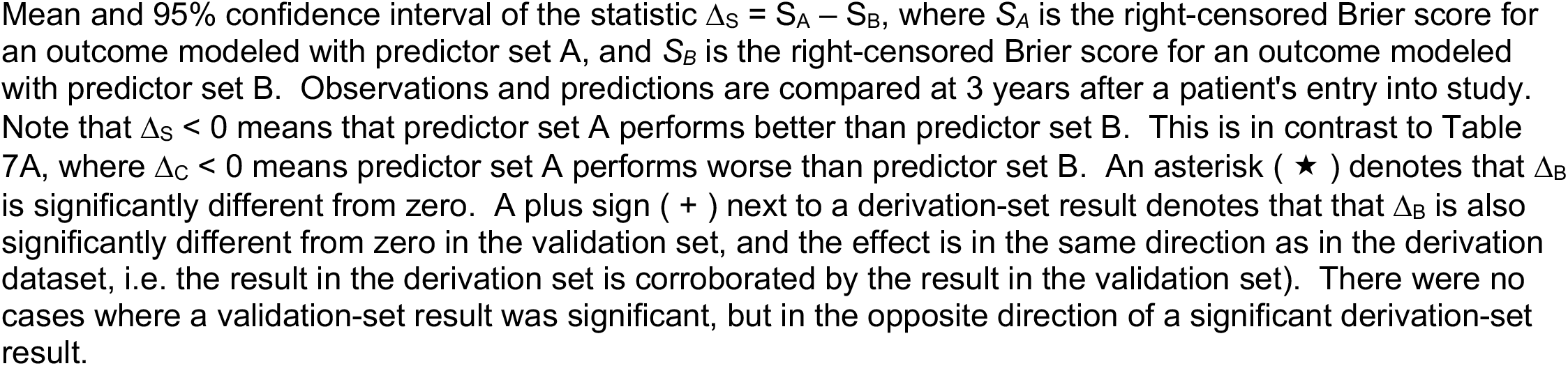
Comparison of pairs of predictor sets according to 3-year Brier Score.

**Supplementary Table 6.**
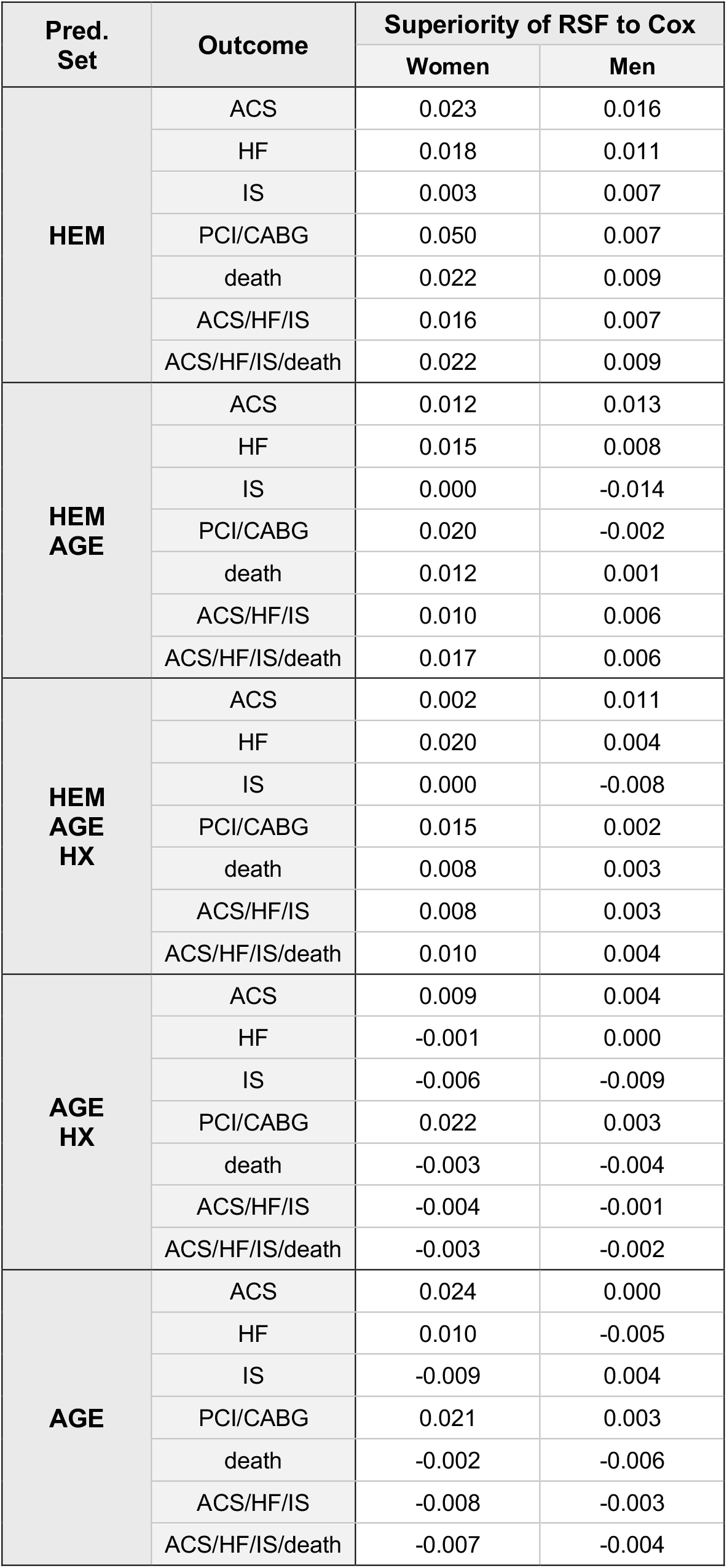
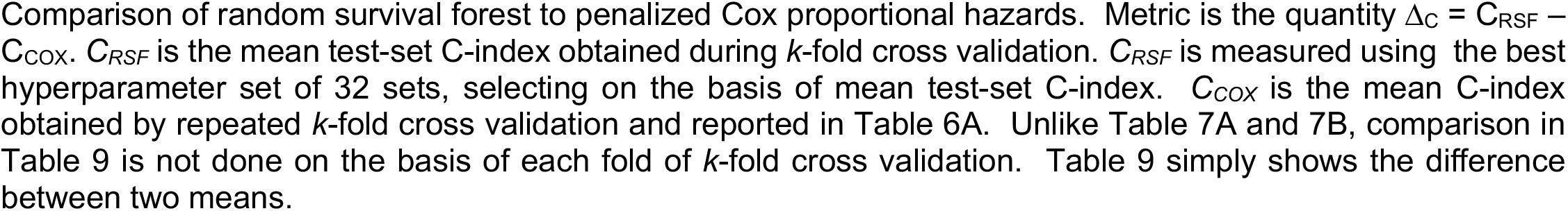
Comparison of random survival forest model to Cox proportional hazards on the basis of C-index.

| Pred. Set | Outcome | Superiority of RSF to Cox |  |
| --- | --- | --- | --- |
|  |  | Women | Men |
| **HEM** | ACS | 0.023 | 0.016 |
|  | HF | 0.018 | 0.011 |
|  | IS | 0.003 | 0.007 |
|  | PCI/CABG | 0.050 | 0.007 |
|  | death | 0.022 | 0.009 |
|  | ACS/HF/IS | 0.016 | 0.007 |
|  | ACS/HF/IS/death | 0.022 | 0.009 |
| **HEM AGE** | ACS | 0.012 | 0.013 |
|  | HF | 0.015 | 0.008 |
|  | IS | 0.000 | -0.014 |
|  | PCI/CABG | 0.020 | -0.002 |
|  | death | 0.012 | 0.001 |
|  | ACS/HF/IS | 0.010 | 0.006 |
|  | ACS/HF/IS/death | 0.017 | 0.006 |
| **HEM AGE HX** | ACS | 0.002 | 0.011 |
|  | HF | 0.020 | 0.004 |
|  | IS | 0.000 | -0.008 |
|  | PCI/CABG | 0.015 | 0.002 |
|  | death | 0.008 | 0.003 |
|  | ACS/HF/IS | 0.008 | 0.003 |
|  | ACS/HF/IS/death | 0.010 | 0.004 |
| **AGE HX** | ACS | 0.009 | 0.004 |
|  | HF | -0.001 | 0.000 |
|  | IS | -0.006 | -0.009 |
|  | PCI/CABG | 0.022 | 0.003 |
|  | death | -0.003 | -0.004 |
|  | ACS/HF/IS | -0.004 | -0.001 |
|  | ACS/HF/IS/death | -0.003 | -0.002 |
| **AGE** | ACS | 0.024 | 0.000 |
|  | HF | 0.010 | -0.005 |
|  | IS | -0.009 | 0.004 |
|  | PCI/CABG | 0.021 | 0.003 |
|  | death | -0.002 | -0.006 |
|  | ACS/HF/IS | -0.008 | -0.003 |
|  | ACS/HF/IS/death | -0.007 | -0.004 |

